# Morphology, Not Motion: Benchmarking Vision-Language Models on Multi-Sign Lung Ultrasound Interpretation

**DOI:** 10.64898/2026.07.23.26358829

**Authors:** Jaeyeon Lee, Athanasios Papastathopoulos-Katsaros, Ilia Buralkin, Zahid Shaik, Brandon Lee, Stephanie Ka-Wing Leung, Michael Alavi, Benjamin Silva, Benjamin Choi, Zhandong Liu, Hyun-Hwan Jeong

## Abstract

**Background:** Point-of-care lung ultrasound (LUS) interpretation requires specialized training. Vision-language models (VLMs) may automate multi-sign assessment but lack systematic clinical benchmarking. We benchmarked three LUS tasks on 125 evaluable cases drawn from 150 point-of-care ultrasound (POCUS) Atlas clips to establish baseline capabilities and isolate clinically relevant failure modes for VLM-based LUS interpretation. **Methods:** We evaluated a normalized multi-model benchmark on three tasks: pleural sliding detection (T1), lung-rocket (B-line) and consolidation classification (T2), and Posterolateral Alveolar and/or Pleural Syndrome (PLAPS; comprising posterolateral consolidation and pleural effusion) assessment (T3). Inputs were 10 uniformly sampled frames for all tasks, plus synthetic M-mode images for sliding. **Results:** We report four findings. First, M-mode improved T1 sliding over static frames for some models, but performance remained modest with wide uncertainty. Second, T2 pathology, identifiable from single frames, was discriminated most clearly above chance on a balanced cohort, with strong lung-rocket and anterior-consolidation F1 achieved by open-weight rather than closed-weight Claude models, although a significant between-model difference held only for lung rockets. Third, T3 PLAPS F1 was high for most models but reflected high positive-class prevalence rather than strong discrimination. Fourth, inter-model agreement was near chance (task-averaged Cohen’s *κ* ≈ 0.02-0.29) and well below each model’s self-consistency, indicating distinct, non-redundant error patterns. **Conclusions:** Static-frame VLMs produced discriminative labels for B-lines and anterior consolidation but remained unreliable for motion-dependent signs (pleural sliding) and prevalence-inflated outcomes (PLAPS). Synthetic M-mode partially recovered temporal information but did not achieve clinically sufficient sliding accuracy. These results support morphology-assisted use for B-lines and consolidation; motion-dependent signs and balanced endpoints require larger validation cohorts before clinical deployment.

## 1 Introduction

Point-of-care ultrasound (POCUS) enables rapid bedside lung assessment, but reliable interpretation requires specialized training that is unevenly distributed across clinical settings. Automating the recognition of individual lung ultrasound (LUS) signs could broaden access, and recent advances in general-purpose vision-language models (VLMs) make this a plausible direction. Achieving this, however, requires understanding where these models succeed and where they fail on real clinical LUS.

Earlier efforts to make this interpretation more reliable have followed two distinct strategies. The first relies on structured clinical frameworks, which give clinicians explicit rules for mapping signs to pathologies during bedside interpretation: the BLUE protocol (**B**edside **L**ung **U**ltrasound in **E**mergency), a rule-based diagnostic algorithm, routes acute respiratory failure through a small set of sign combinations with a reported diagnostic accuracy of 90.5% (Lichtenstein and Mezière 2008), and international consensus recommendations standardize the underlying sign vocabulary across emergency and critical care settings (Volpicelli et al. 2012). More recent variants of this framework-based approach, such as the Pleura-ABCDE taxonomy, extend its logic to neonatal lung ultrasound (Sandig et al. 2025). The second strategy is computational: such approaches have, in parallel, targeted single tasks such as B-line detection and pleural effusion classification using supervised learning on curated datasets (Born et al. 2020; Roy et al. 2020). These systems can be accurate within their scope, but each addresses one binary question, and none captures the multi-sign, multi-pathology reasoning that clinical LUS interpretation demands. General-purpose VLMs—models trained on broad non-medical image and text data rather than fine-tuned for ultrasound—including both open-weight families such as Gemma, MedGemma, and Qwen and frontier closed-weight systems such as Anthropic Claude, are, like the early GPT-4V demonstrations (Yang et al. 2023), notable because they can produce structured, multi-sign output in a single inference pass without task-specific fine-tuning, collapsing what previously required several specialized models into one prompt.

Applying these models to LUS, however, exposes a fundamental mismatch. LUS is inherently a video examination: several of its most important signs are defined by motion and have no single-frame signature. The clearest example is lung sliding, the shimmering sonographic appearance of the pleura sliding to and fro during respiration that excludes pneumothorax. Most VLMs, in contrast, ingest still images or a handful of sparsely sampled frames. Native video understanding is a recent and still-limited capability. Video-capable LLMs as a class are an emerging research area (Tang et al. 2024) and are claimed by only a small number of models. General-domain video benchmarks—those built from everyday, non-medical video rather than clinical imaging—that isolate temporal reasoning, most directly the 20-task MVBench evaluation, find that current multimodal models remain far from satisfactory on tasks that cannot be solved from a single frame (Li et al. 2024). LUS-specific temporal evaluation is lacking, leaving it unclear whether that gap transfers to bedside LUS, and creating uncertainty about how it does so. As a result, several frontier video-capable models have not been evaluated on LUS at all, and the field still lacks a clear picture of how the static-versus-temporal divide shapes performance on clinically structured LUS tasks.

The evaluation landscape compounds this gap. Recent work has begun to benchmark VLMs on ultrasound understanding, most notably U2-BENCH, which spans fifteen anatomical regions including the lung (Le et al. 2026), and dedicated single-organ efforts such as FETAL-GAUGE for fetal imaging (Alasmawi, Saeed, and Yaqub 2025). Each, however, leaves the lung-ultrasound question largely open. U2-BENCH evaluates models only on static images and image-level tasks such as classification, view recognition, and report generation, so it does not probe the multi-frame or temporal understanding that motion-dependent signs require, and the lung is just one of fifteen regions rather than a task-structured target. FETAL-GAUGE is likewise image-based and dedicated to fetal rather than pulmonary imaging. Closest in spirit, COVID-BLUeS prospectively studied AI for lung ultrasound but was scoped narrowly to COVID-19 detection and severity in a small cohort, and its authors explicitly identify frame-based processing that ignores video-level information, together with dataset heterogeneity, as key factors limiting model performance (Wiedemann et al. 2025). The contemporaneous Stanford OpenPOCUS database (Kumar et al. 2026) expands the public landscape with sign-level video annotations for B-lines, consolidation, indeterminate, and no-abnormality across anterior, lateral, and posterior zones. However, its multi-center emergency-department cohort, in which roughly half of the cases were COVID-19 pneumonia, does not annotate sliding or lung-point findings, nor does it distinguish pleural effusion and Posterolateral Alveolar and/or Pleural Syndrome (PLAPS)-specific patterns. As such, it does not independently support the BLUE-style sign hierarchy that our benchmark targets. Two gaps therefore remain. First, frontier video-capable VLMs have not been evaluated on LUS. Second, no public benchmark covers the full lung-ultrasound sign vocabulary that BLUE-protocol bedside interpretation traverses: sliding and lung-point at the pleural line, B-lines and consolidation, and pleural effusion. Model performance on LUS has therefore been measured only on partial slices of the clinical decision tree.

We address these gaps with a three-task benchmark on 125 evaluable cases drawn from 150 POCUS Atlas clips: sliding detection (T1), lung-rocket/B-line and consolidation classification (T2), and PLAPS assessment (T3). The three tasks were chosen to cover the BLUE protocol’s lung sign-level vocabulary on a single cohort, allowing T1 (motion-dependent) and T2/T3 (morphology-dependent) findings to be compared against the same models. Every headline metric is summarized as the mean F1 across five complete runs per model with a 95% confidence interval, and between-model ranking claims are supported by a prespecified paired comparison, distinguishing findings that reflect stable model behavior from those driven by single-run noise. Together these design choices establish baseline capabilities and isolate the failure modes that matter for VLM-based LUS interpretation.

## 2 Methods

### 2.1 Dataset

The evaluation dataset comprises 150 LUS video clips sourced from The POCUS Atlas^1^, an open-access point-of-care ultrasound education repository. Cases span two atlas partitions: the adult lung collection (116 cases) and the pediatric lung collection (34 cases). Each case includes a descriptive title, contributor-assigned categories and tags, a free-text clinical vignette, and a short MP4 video clip. The dataset is publicly hosted on HuggingFace under a CC-BY-NC-4.0 license. POCUS Atlas was selected as the open LUS video source whose scope and metadata best support the multi-sign, multi-task evaluation our benchmark requires; positioning against other public LUS video datasets is given in Section 1. The per-task adult-to-pediatric breakdown and a descriptive per-source F1 decomposition are reported in Appendix D.

The 125 benchmark-eligible cases (Fig. 1) constitute the evaluation set and are distributed across three evaluation tasks (T1, T2, T3) by anatomical scan region, with case counts and class balance given in Table A4 and per-case exclusion reasons itemized in Table A5.

**Figure 1:**
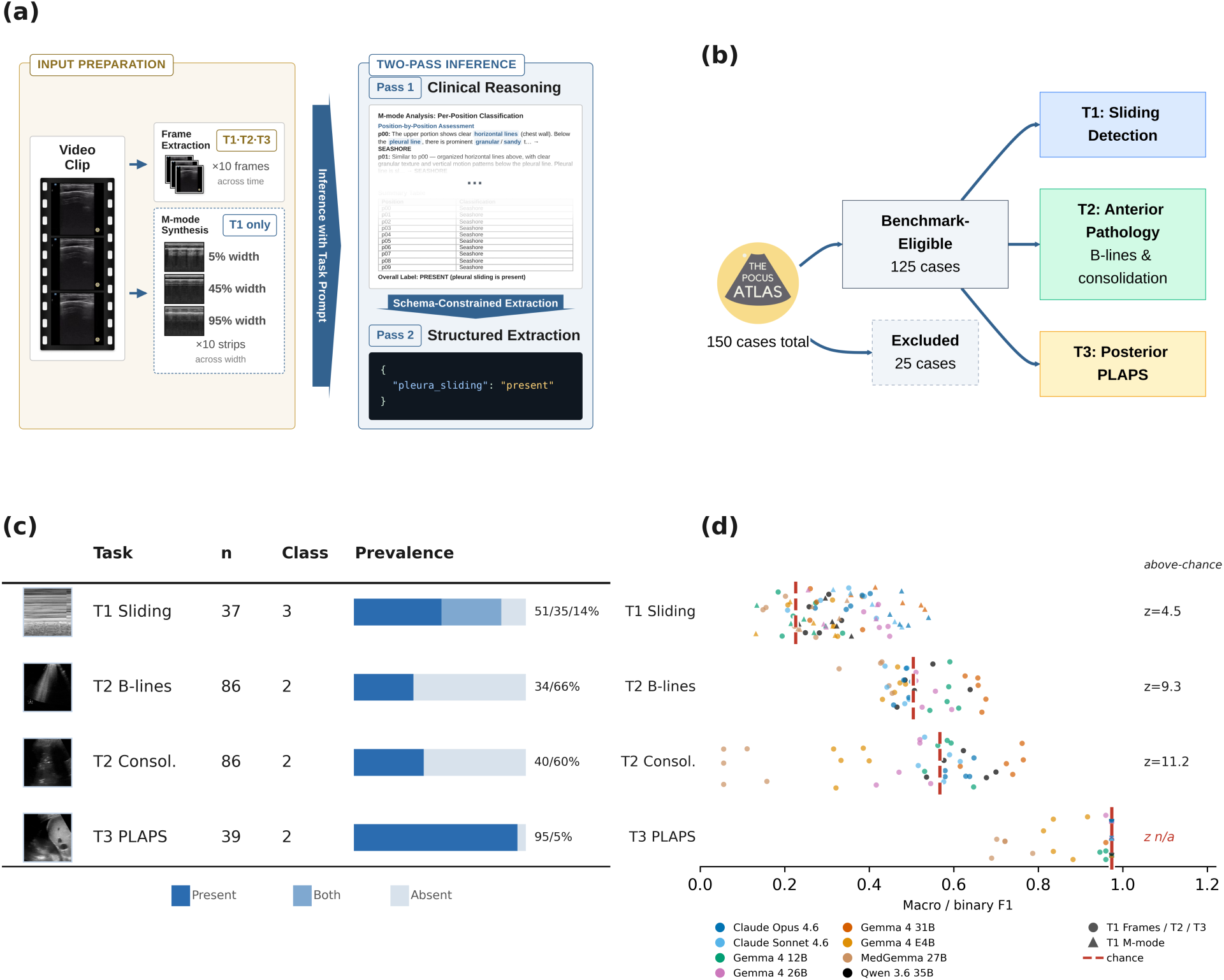
POCUS multi-task LUS benchmark overview. (a) Two-pass inference pipeline: input preparation produces 10 uniformly-sampled frames (all tasks) or 10 synthetic M-mode strips (T1 only); Pass 1 generates free-text clinical reasoning and Pass 2 extracts structured labels into a per-task JSON schema. (b) Dataset partitioning: of 150 total POCUS Atlas cases, 25 are excluded (non-pulmonary content, single-frame videos, duplicates, non-diagnostic), leaving 125 benchmark- eligible cases assigned to three evaluation tasks by anatomical scan region; 36 sliding-assessed cases appear in both T1 and T2. (c) Per-task dataset statistics (case count and class balance); exact values are tabulated in Table A4. (d) Results summary: per model-run F1 (points, colored by model) for each task against the label-shuffle permutation chance baseline (dashed line), with the above-chance standardized effect size (z; higher z means the model discriminates further above random-guess chance) labeled per task; T3 PLAPS’s F1 cloud sits on its chance line (z not defined), indicating near-chance discrimination (see Appendix F). In-figure task labels are abbreviated: “T2 Consol.” denotes T2 consolidation and “T3 PLAPS” denotes the T3 posterolateral alveolar and/or pleural syndrome (PLAPS) task.

### 2.2 Annotation

Lung-ultrasound signs are identified by their sonographic appearance or motion rather than by the scanning location at which they are seen. Within the BLUE protocol, however, the anterior versus posterolateral distinction is part of how signs map to pathology, as the anterior C-profile and the posterolateral PLAPS are location-defined (Lichtenstein and Mezière 2008).

Ground-truth annotations were derived in two stages by applying regex patterns to the concatenation of each case’s title, categories, and tags. The free-form clinical vignette was excluded to prevent spurious matches. First, pattern matching produced per-feature labels Table 1. Second, these features were combined to classify each case as *anterior* or *posterior* : cases with PLAPS or normal-posterior markers (excluding pneumothorax) were classified as posterior. Cases with lung consolidation, lung-point, assessed sliding, B-lines without PLAPS, or A-line findings, and no posterior markers, were classified as anterior. This classification determined task eligibility (T2: anterior; T3: posterior), while T1 included all anterior cases with an assessed sliding label. This zone partition follows the BLUE-protocol scan stages (Lichtenstein and Mezière 2008): because B-profiles (lung rockets/B-lines) and C-profiles (anterior consolidation) are determined at the stage-1 anterior upper/lower BLUE points, whereas posterolateral consolidation and pleural effusion are captured separately as PLAPS at the stage-3 posterolateral point, we define T2 as an anterior-zone task and T3 as the posterolateral PLAPS task. This assignment is derived from annotation text, not from recorded probe position (which the POCUS Atlas does not provide), and therefore represents an inferred rather than a measured scan zone. This regex-based approach may miss atypical phrasing or incorrectly resolve negation in borderline cases. These limitations are discussed in Section 5.

**Table 1:** Ground-truth annotation rules (selected examples).

| Label | Pattern examples | Type |
| --- | --- | --- |
| Sliding | “lung slid”<sup>2</sup>, “seashore” | 3-class |
| B-lines | “b-lines”, “interstitial”, “confluent” | bool |
| Consolidation | “consolidation”, “hepatization”, “shred sign” | bool |
| PLAPS | “pleural effusion”, “spine sign”, “jellyfish” | bool |

Labels are derived by regex matching against concatenated case metadata. Sliding is 3-class: *present* if affirmative patterns remain after negation removal, *absent* if negation or pneumothorax terms match, and *both* if “lung point” or co-occurrence. The complete pattern set is given in Table A1 and Table A2.

The LUS signs we evaluate differ in the information needed to detect them. Lung sliding is a temporal sign, the shimmering to-and-fro movement of the pleural line across the respiratory cycle, and its loss raises concern for pneumothorax. Because sliding is defined by motion, it can be judged only from successive frames. The remaining signs are morphologic and are read from a single frame. A-lines, B-lines, and consolidation form an aeration continuum that reflects progressive loss of parenchymal air (Volpicelli et al. 2012), whereas pleural effusion is a separate pleural-space finding captured within PLAPS. This distinction shapes our single-image evaluation. A still frame cannot convey the motion that sliding requires, so we encode sliding as a synthetic M-mode image in T1 and evaluate the morphologic signs from single frames in T2 and T3.

### 2.3 Evaluation Tasks

The three evaluation tasks are organized around the BLUE protocol (Lichtenstein and Mezière 2008). T1, T2, and T3 collectively cover all lung sign-level questions in the protocol; only the integration step (combining signs into a profile and pathology diagnosis) and the venous deep vein thrombosis assessment (a lower-extremity vascular scan separate from lung imaging) remain outside scope. An appendix figure maps each task to the protocol’s sign-recognition nodes (Fig. E1).

**T1 - Sliding Detection** performs three-way classification (present / absent / both) over 37 sliding-assessed cases. *Both* indicates a within-clip transition between sliding and absent sliding and signifies the lung-point sign, the transition point where the pleural line shifts from sliding to non-sliding across the field of view as the probe traverses the boundary of a pneumothorax. The ground-truth distribution comprises present (19; 51%), both (13; 35%), and absent (5; 14%). **T2 - Anterior Pathology** consists of two binary classification subtasks on 86 anterior-window cases: (a) lung rocket/B-line detection (present/absent, prevalence 34%), and (b) consolidation detection (present/absent, prevalence 40%). Consolidation sub-types (air bronchogram, shred sign) were recorded but not used as primary metrics. **T3 - Posterior PLAPS** performs binary classification (pathological PLAPS present/absent) on 39 posterior-window cases (prevalence 95% positive; 37 of 39 cases), reflecting that the majority of posterior cases in the POCUS Atlas demonstrate pleural effusion and/or consolidation.

Case-to-task assignment was performed programmatically via multi-column derivation from the annotation matrix: T1 includes all cases with a non-missing sliding assessment; T2 selects anterior- zone cases using a composite of consolidation, lung-point, sliding, B-line, and A-line annotations while excluding posterior cases; T3 selects cases positive for PLAPS or normal posterior findings while excluding pneumothorax. Task sets are not mutually exclusive as sliding-assessed anterior cases appear in both T1 and T2.

### 2.4 Data Processing

Video frames were extracted using uniform temporal sampling: for each clip, 10 evenly-spaced frames were selected via linear interpolation across the total frame count.

For the T1 sliding detection task, synthetic M-mode images were generated to encode temporal dynamics in a spatial representation amenable to single-image VLM input. The algorithm proceeds as follows: (1) all video frames are converted to grayscale; (2) the active ultrasound region is identified via temporal standard deviation thresholding (15% of peak variance), excluding static mask borders; (3) 10 lateral positions are sampled at 5%, 15%, . . ., 95% of the active region width; (4) at each position, a 20-pixel-wide vertical strip is averaged across the strip width to produce one column per frame, then stacked temporally to form the M-mode image; (5) contrast-limited adaptive histogram equalization (CLAHE, clip limit 3.0) is applied; and (6) the result is resized to a 4:3 landscape aspect ratio. Three cases with spliced display panels required manual vertical crop overrides.

### 2.5 Pipeline Architecture

The inference system enforces a uniform output schema across models through a two-pass pipeline (Fig. 1 (a)): Pass 1 generates free-text clinical reasoning over the preprocessed input images (Section 2.4), and Pass 2 extracts structured labels from the reasoning trace into a predefined JSON schema. Pass 2 receives only the text reasoning from Pass 1 (no images) together with the extraction schema. Schema enforcement is implemented via forced tool-use (Anthropic API) or structured output mode (OpenAI-compatible endpoints, used both for hosted-API models and for locally-served GGUF deployments), both of which constrain token generation to valid JSON conforming to the task schema. The benchmark covers eight models spanning open-weight Gemma and Qwen families, the medical-domain MedGemma variant, and Anthropic Claude Sonnet and Opus; per-model identifiers, providers, and parameter counts are listed in Table C1. Task-specific prompt templates were designed for T1 (sliding assessment), T2 (lung rockets/B-lines + anterior consolidation), and T3 (posterior PLAPS).

### 2.6 Metrics

The F1 score (or F-measure) is the harmonic mean of precision and recall, a standard summary metric for classification that penalizes both false positives and false negatives in equal proportion. We adopt it as the primary task metric: macro-averaged F1 for the multi-class T1 task and binary F1 for T2 and T3. F1 was chosen because, on tasks where false positives and false negatives carry different clinical costs, it captures both precision and recall on the positive class with a single number, in contrast to plain accuracy, which is dominated by the majority class on imbalanced cohorts (notably T3). For a given class, precision and recall are defined from the counts of true positives (*TP*), false positives (*FP*), and false negatives (*FN*):

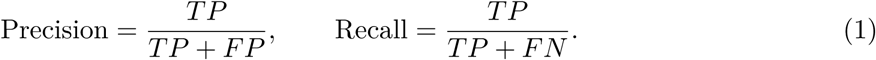

The F1 score is the harmonic mean of precision and recall:

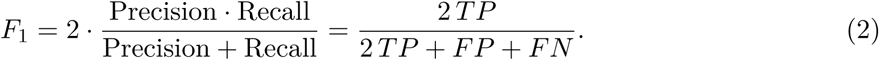

For binary tasks (T2, T3) F1 is computed on the positive class. For the multi-class T1 task, the macro-averaged F1 averages the per-class F1 scores with equal weight across the *C* classes (present, absent, both), so that minority classes are not dominated by prevalence:

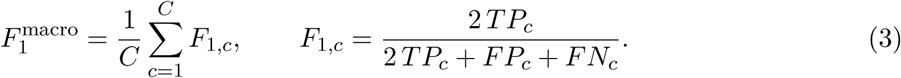

All eight benchmark models were evaluated over *N* = 5 independent complete runs per task (see Table C1).

Confidence intervals shown in the headline F1 figures are 95% confidence intervals across each model’s five complete runs (t-distribution, four degrees of freedom), and the plotted point estimate is the mean F1 over those runs. Confusion matrices summarize a single representative complete run per model and are used to interpret whether high F1 reflects true discrimination or class-prevalence artifacts.

We apply a between-model significance test only where the manuscript makes a model-ranking claim, which is limited to T2: T1 performance is interpreted as a input-representation effect rather than a model ranking (Section 3.2), and T3 F1 is a cohort-prevalence artifact rather than a measure of discrimination (Section 3.4), so neither supports a between-model comparison. Where a ranking is claimed, we test the two highest-mean-F1 models using per-case correctness on the shared cases. Runs are paired chronologically, and we report an exact McNemar test on the pooled discordant pairs (a binomial test on the counts of cases where one model is correct and the other is wrong) together with a paired t-test on the five per-run F1 values. A difference is described as established only when both tests reject at the 0.05 level; otherwise the ranking is reported descriptively.

To assess whether the best model on each task performs above chance, we compare its observed F1 against a label-shuffle null. Holding the model’s predictions fixed, we permute the ground-truth labels within each complete run, recompute the same mean-F1 statistic across runs, and repeat this 10{,}000 times to build a null distribution. We summarize the result with a standardized effect size *z*, the number of null standard deviations separating the observed F1 from the mean of the shuffle distribution, together with the reference chance F1 of a no-information predictor (an all-positive predictor, 2*p/*(1 + *p*), for the binary tasks, and the majority-class predictor macro-F1 for the multi-class T1 task). This permutation-based above-chance analysis is a separate procedure from the bootstrap resampling used for the across-run confidence intervals in the headline rankings: the permutation null asks whether performance exceeds chance, whereas the bootstrap CIs quantify run-to-run variability. When the permutation null is degenerate (all permutations return the identical F1, so its variance is zero), *z* is undefined; this occurs on T3, where the best model is all-positive (see Section 3.4). Per-task above-chance results are reported in Appendix F.

## 3 Results

### 3.1 Overview

Across the three tasks (summarized in Fig. 1 (d)), models performed best when a finding could be identified from a single still image. Performance degraded when the finding depended on motion, and it became uninterpretable when the cohort was insufficiently balanced to separate classification skill from chance-level performance. For anterior morphology (T2), detection of consolidation and lung rockets/B-lines reached mean binary F1 scores of 0.721 and 0.657 respectively. Because the cohort is balanced, these values reflect genuine discrimination rather than a prevalence artifact. Pleural sliding (T1) was the most difficult task. Static frames left the best model at only ∼0.42 macro-F1, and converting the clips into synthetic M-mode lifted the ceiling only to ∼0.49, with wide confidence intervals. Posterior PLAPS (T3) showed high apparent F1 (up to 0.974), but this reflects an overwhelmingly positive cohort rather than discriminative ability. Its raw F1 is therefore inflated relative to T2’s balanced-cohort F1 and cannot be ranked against it. Inter-model concordance was near chance and well below within-model self-consistency, indicating that errors were not systematically shared across models.

The following sections outline each task, highlight where the models succeeded and where they failed, and analyze why some findings are more recoverable from sparse-frame input than others.

### 3.2 T1: M-mode Partially Improves Sliding Detection

T1 is best interpreted by comparing input representations before comparing models. With static frames, every model performed poorly, the best condition reaching only 0.425 macro-F1 (Gemma 4 26B A4B; Fig. 2). Recasting the same clips as synthetic M-mode images raised the ceiling to 0.486 (Claude Opus 4.6), a real but partial improvement that remains inadequate for clinical sliding classification and whose confidence intervals overlap heavily with the static-frame results.

**Figure 2:**
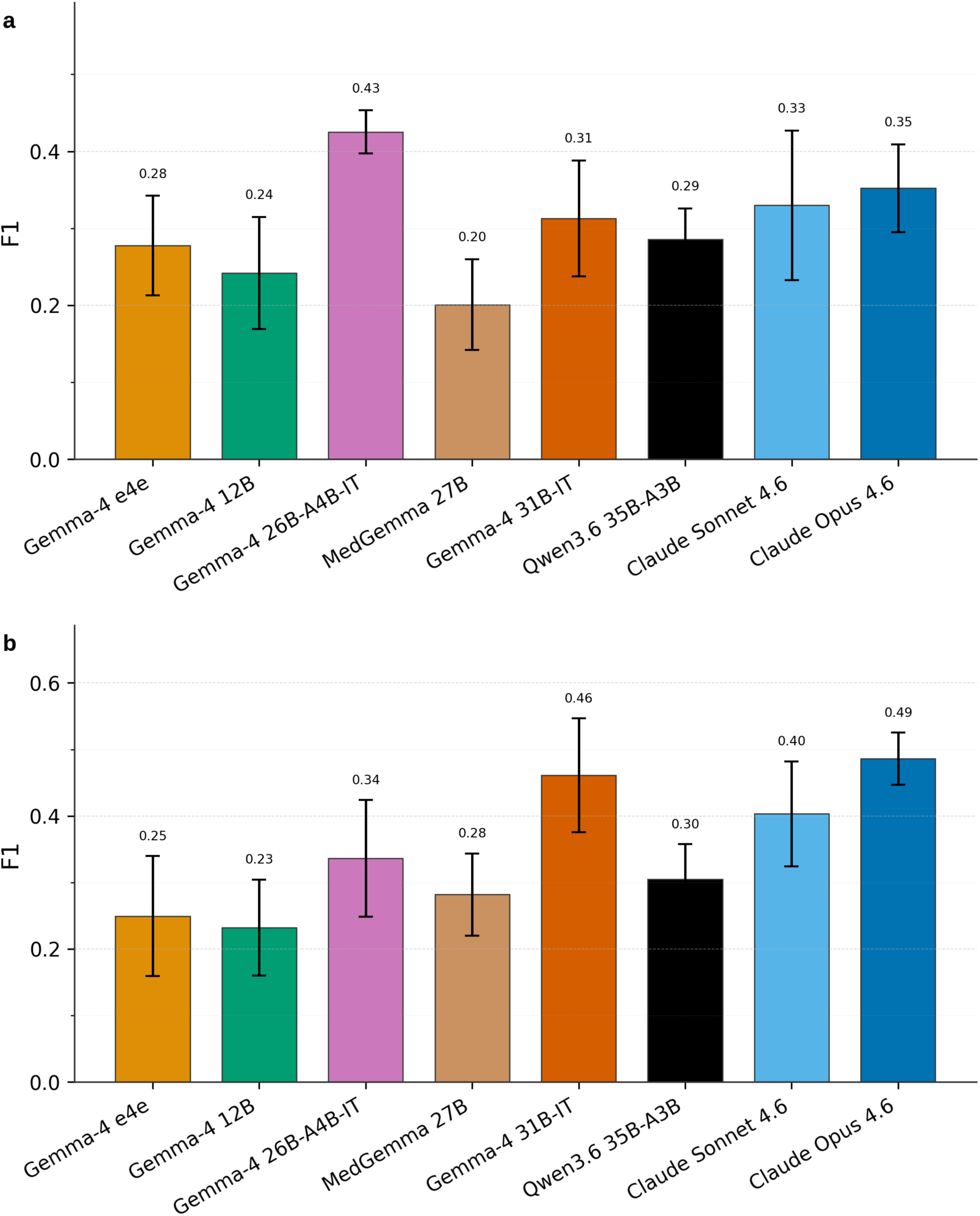
T1 pleural sliding macro-F1 (mean over five runs per model, with error bars showing 95% confidence intervals across runs). (a) Static-frame input: the best plotted condition reaches 0.425 macro-F1 (Gemma 4 26B A4B). (b) Synthetic M-mode input: the strongest condition shifts to Claude Opus 4.6 (F1 0.486), but performance remains modest and uncertainty is substantial.

The confusion matrices (Fig. 3) explain why this task is difficult. Lung sliding is a motion sign: a clinician recognizes it from the shimmer of the pleural line over time, not from any single frame. Ten still frames distribute that temporal cue across images that individually appear almost identical whether sliding is present or absent, so the models predict the majority *present* class. Synthetic M-mode helps because it encodes the temporal information into a single spatial image, thereby generating the seashore–barcode contrast used to distinguish lung sliding. However, it does not fully restore the signal a clinician would obtain from the live scan. Two factors compound the difficulty. The *absent* class contains only five ground-truth cases, so a few misclassifications swing the score.

**Figure 3:**
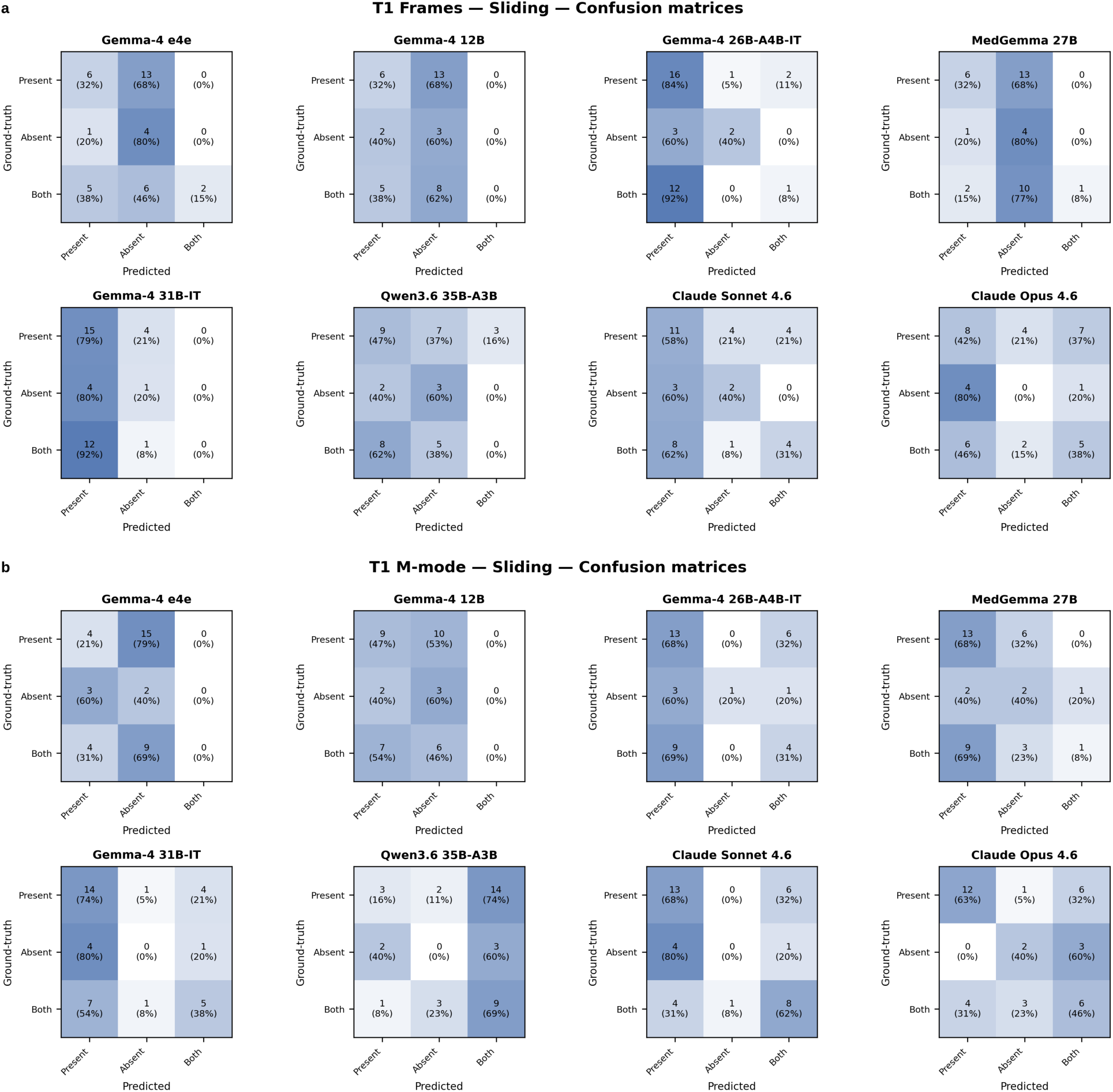
T1 confusion matrices for (a) static-frame input and (b) synthetic M-mode input. Errors concentrate in the underpowered absent class (only five ground-truth cases) and in the lung-point “both” cases, where sliding transitions within the scan.

The lung-point “both” cases also require the model to notice that sliding *changes* across the field of view — a within-clip transition that is the easiest cue to lose when frames are sampled independently.

T1 is therefore the benchmark’s primary failure mode, with the cause being representational rather than model-capability-limited. Both T1 variants exceed the majority-class chance macro-F1 of 0.226 (best macro-F1 0.425 for frames and 0.486 for M-mode, standardized effect *z* of 4.5 against the label-shuffle null). The best model therefore discriminates above chance, but the absolute F1 stays low.

### 3.3 T2: Anterior Pathology Is Discriminated Above Chance from a Single Frame

Both anterior findings can be identified from a single frame, and the cohort is balanced, so T2 F1 measures discrimination rather than prevalence (Fig. 4). T2 is the task on which a model most clearly reads clinically meaningful morphology, with the best model exceeding the prevalence-implied chance F1 on both findings. For lung rockets/B-lines, Gemma 4 31B achieved the strongest F1 (0.657) and its lead over the next model (Gemma 4 12B) is supported by a paired comparison. The pooled exact McNemar test is significant (p < 0.001) and the paired t-test on per-run F1 agrees (t = 4.32, p = 0.012). For consolidation, the leading open-weight models cluster closely (Gemma 4 31B, Qwen3.6 35B A3B, and Claude Opus 4.6 all near 0.62–0.72) and no model is statistically separable from the others (McNemar p = 0.18, paired t p = 0.12), so we make no leader claim for consolidation. The full paired-test statistics are given in Table G1.

**Figure 4:**
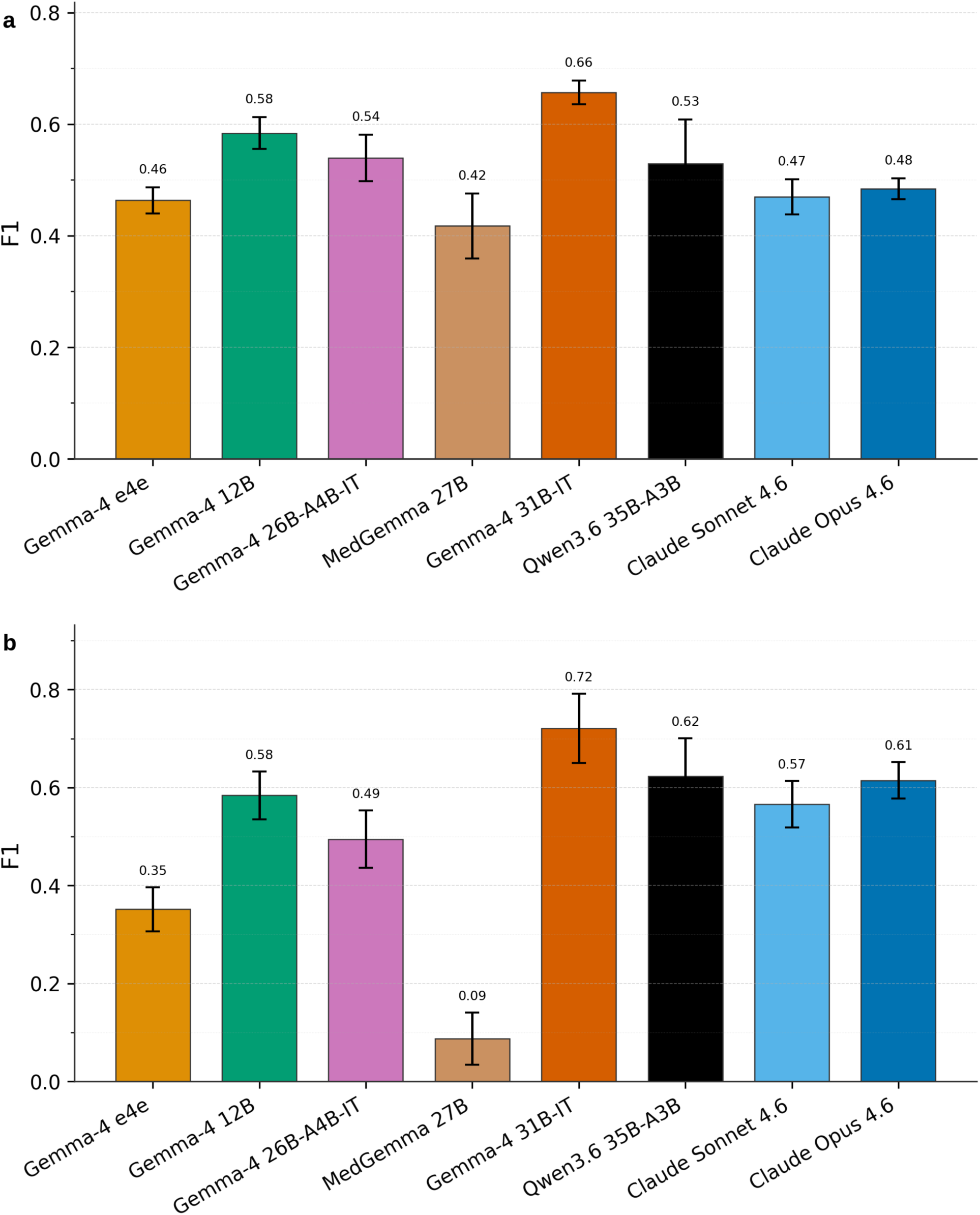
T2 anterior pathology F1 (mean over five runs per model, with error bars showing 95% confidence intervals across runs). (a) Lung rockets/B-lines: detection peaks with gemma-4-31b-it (F1 0.657). (b) Consolidation: detection peaks with gemma-4-31b-it (F1 0.721).

Anterior pathology falls on the benchmark’s static-frame side: both findings are encoded in the texture of a single frame. B-lines are bright, vertical comet-tail artifacts arising at the pleural line; consolidation appears as tissue-like, hepatized lung with recognizable internal structure. Neither sign needs motion to be identified. Unlike T1, which loses information when frames are sampled, these signs remain fully visible in individual frames. The confusion matrices (Fig. 5) show that errors are distributed across both classes rather than collapsing onto one, which is why F1 here measures discrimination instead of prevalence.

**Figure 5:**
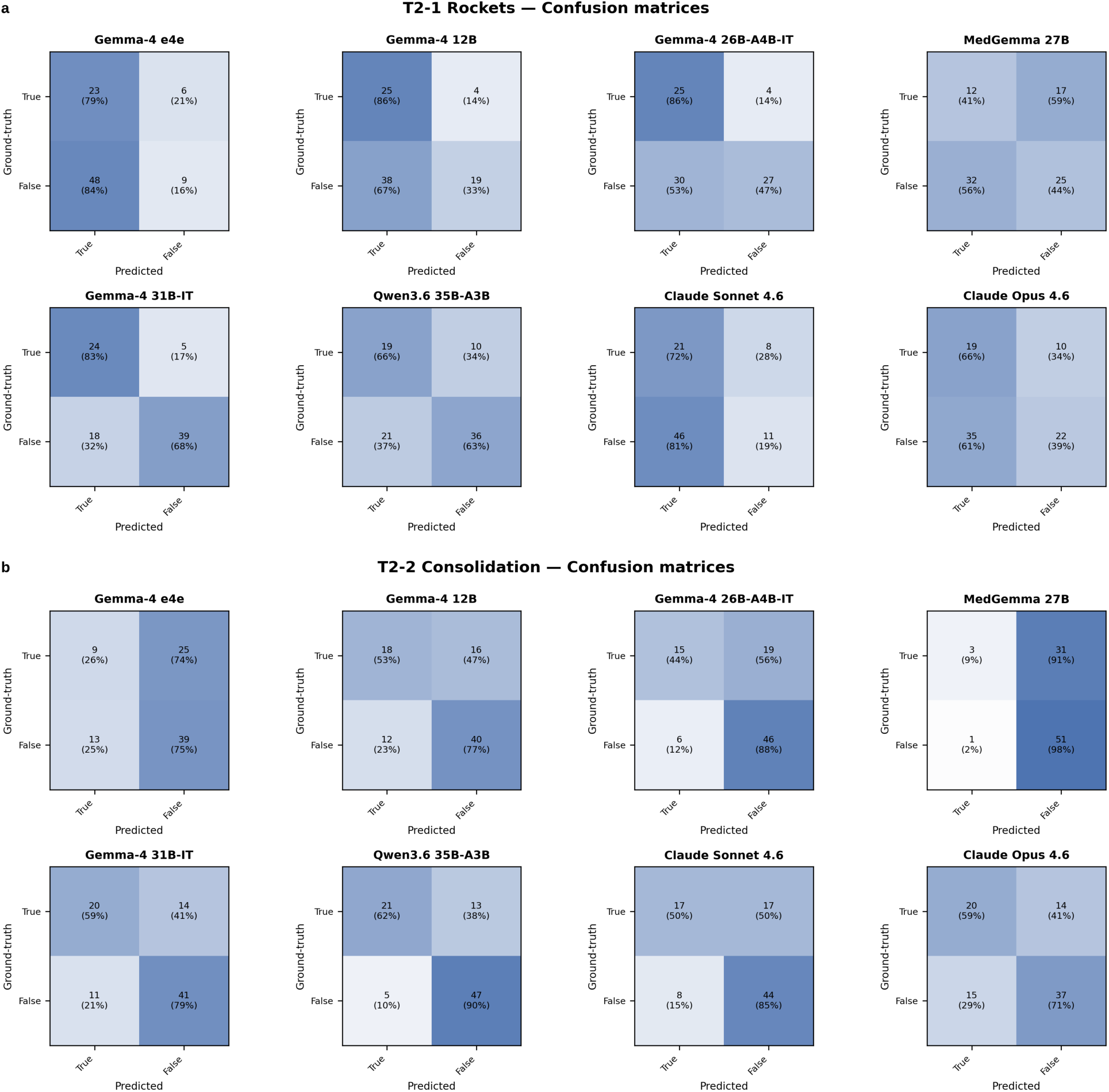
T2 confusion matrices for (a) lung rockets/B-lines and (b) consolidation. Errors are distributed across both classes, indicating that F1 reflects genuine discrimination rather than prevalence.

A label-shuffle permutation test confirms that both findings clear the prevalence-implied chance F1 by a substantial margin. Lung rockets/B-lines reached 0.657 against a chance F1 of 0.504, and consolidation reached 0.721 against 0.567, each sitting far above the shuffle null (standardized effect *z* of 9.3 and 11.2). T2 is the only task where the best model exceeds chance by this much, which is what makes it the benchmark’s strongest evidence of genuine discrimination rather than a prevalence artifact. The full per-task comparison is given in Table F1.

### 3.4 T3: High PLAPS F1 Is Not Evidence of Specificity

Several models reached F1 ∼0.97 on PLAPS (Fig. 6), but this score reflects cohort prevalence rather than discriminative ability. The confusion matrices (Fig. 7) show that this score does not reflect specificity: the posterior cohort splits 37 positive to 2 negative, and the high-scoring models predicted positive for nearly all cases. That strategy yields perfect sensitivity and a high F1 while contributing near-zero specificity: the two negative cases are not reliably identifiable. The score is therefore prevalence-saturated: it rewards a model for matching the base rate, not for discriminating effusion from normal posterior anatomy. MedGemma-27B was the only model to correctly classify both negative cases (TN = 2), but its low recall (recall *→* 0.54) suggests that this may reflect a general tendency to predict negative rather than reliable discrimination. Because the evaluation included only two negative cases, the result should be interpreted cautiously.

**Figure 6:**
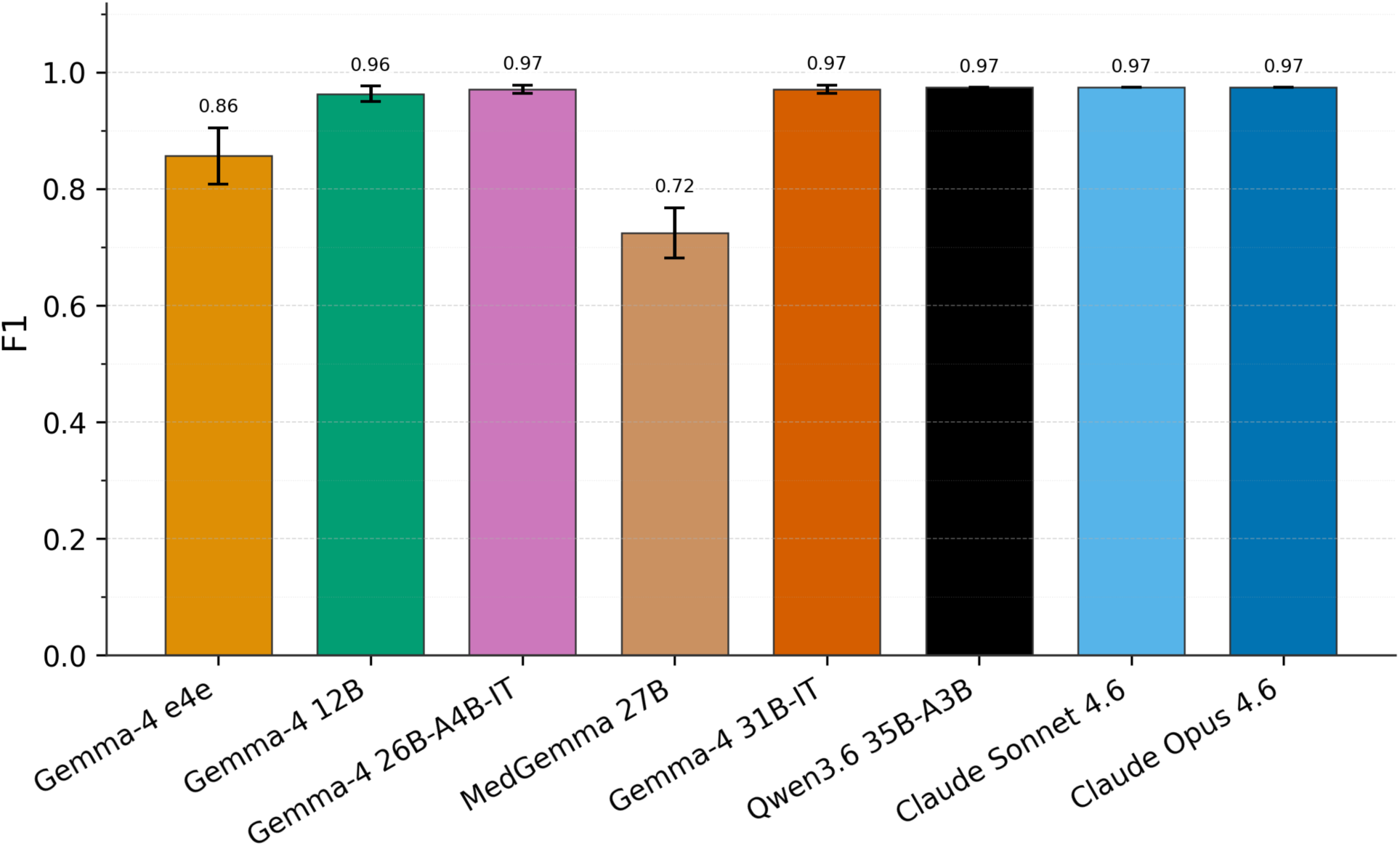
T3 posterior PLAPS F1. Several models cluster near 0.974, but this high score must be interpreted against the highly imbalanced ground truth.

**Figure 7:**
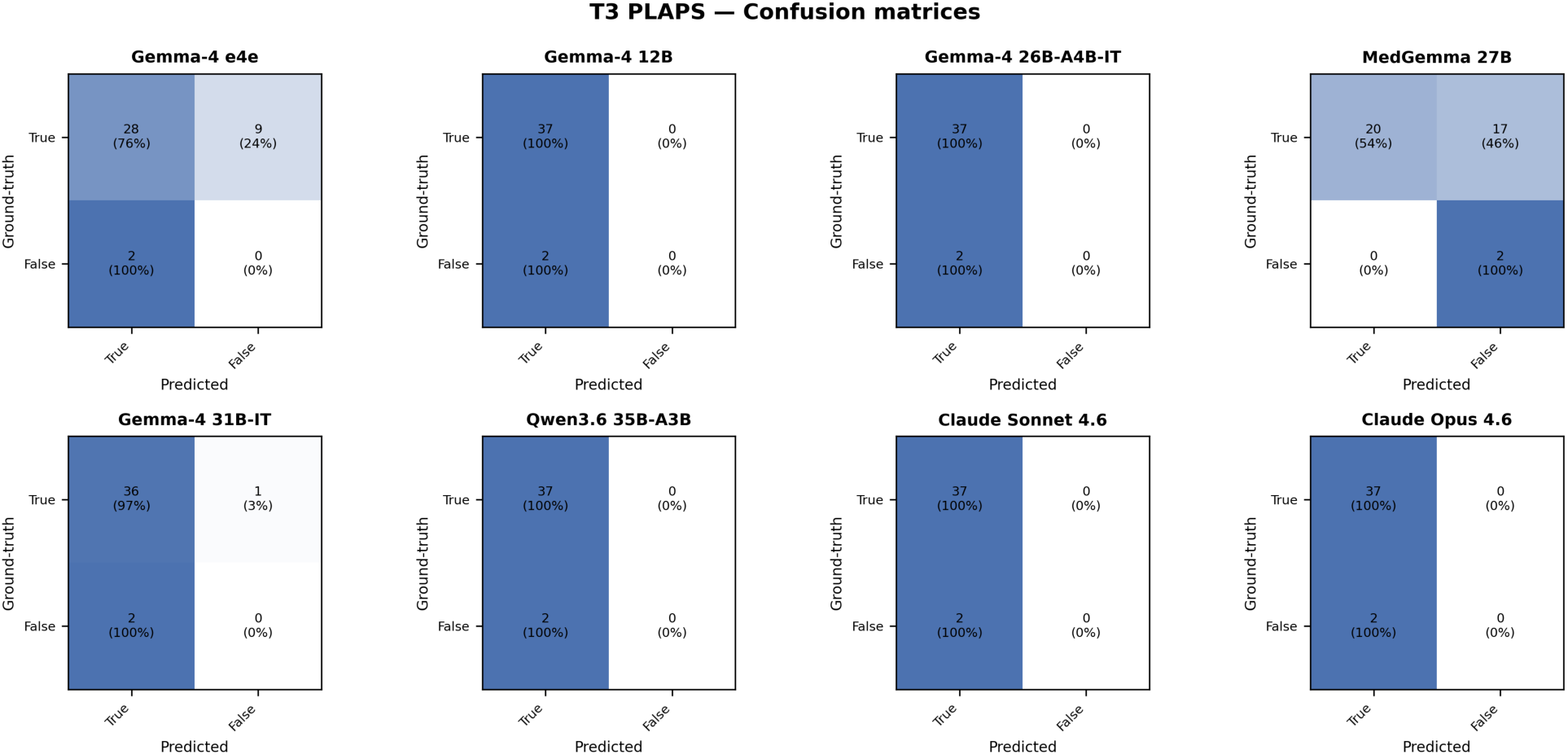
T3 PLAPS confusion matrices. Most high-F1 models predict all posterior cases as positive, yielding perfect sensitivity but zero specificity on the negative cases.

The factor limiting evaluation is the cohort composition rather than the difficulty of the imagery. Distinguishing a true PLAPS from normal posterior findings is itself nontrivial (lung curtain and other normal artifacts can mimic effusion-like appearances when temporal context is limited), but with only two negatives we cannot measure whether any model handles that distinction. Until the posterior cohort includes substantially more negative cases, T3 F1 should be read as a dataset- balance warning rather than as evidence of posterior competence. A label-shuffle null (a permutation test, not the bootstrap used for the across-run confidence intervals) makes this explicit. The best model, Claude Opus 4.6, predicted positive for every posterior case in all five runs. For such an all-positive predictor the F1 depends only on how many cases are positive, and shuffling the ground-truth labels does not change that count; every permutation therefore returns the identical F1, so the null collapses to a single value with zero spread (zero variance). Its F1 (0.974) sits exactly on this degenerate chance value, leaving no separation to standardize and *z* undefined: the observed 0.974 is indistinguishable from a no-information baseline on this cohort.

### 3.5 Pairwise Model Agreement and Error Diversity

This section evaluates whether the eight models behave as interchangeable predictors or retain distinct error patterns, by comparing inter-model agreement against per-model self-consistency (Fig. 8). Self-consistency, reported on the diagonal, characterizes the stochastic variability of each model across five repeated runs and provides a within-model reference for interpreting inter-model agreement. Self-consistency varies substantially across the benchmark: Claude Opus and Claude Sonnet reach 0.64 and 0.51, respectively; the remaining models lie between 0.13 and 0.36, with Gemma-4-31B (0.36), Qwen3.6-35B (0.31), and Gemma-4-26B (0.30) clustered near the upper end of that range, Gemma-4-12B (0.26) and MedGemma-27B (0.18) below, and Gemma-4-e4e at 0.13. Off-diagonal *κ* values range from -0.01 to 0.29 and are lower than the corresponding diagonals for every model. The largest inter-model *κ*, between Claude Opus and Claude Sonnet at 0.29, remains below either Claude model’s self-consistency (0.64 and 0.51), and the remaining inter-model values lie between 0.02 and 0.17. One model pair (Gemma-4-31B with MedGemma-27B) yields slightly negative *κ*, reflecting marginal above-chance disagreement on at least one task rather than a systematic adversarial relationship.

**Figure 8:**
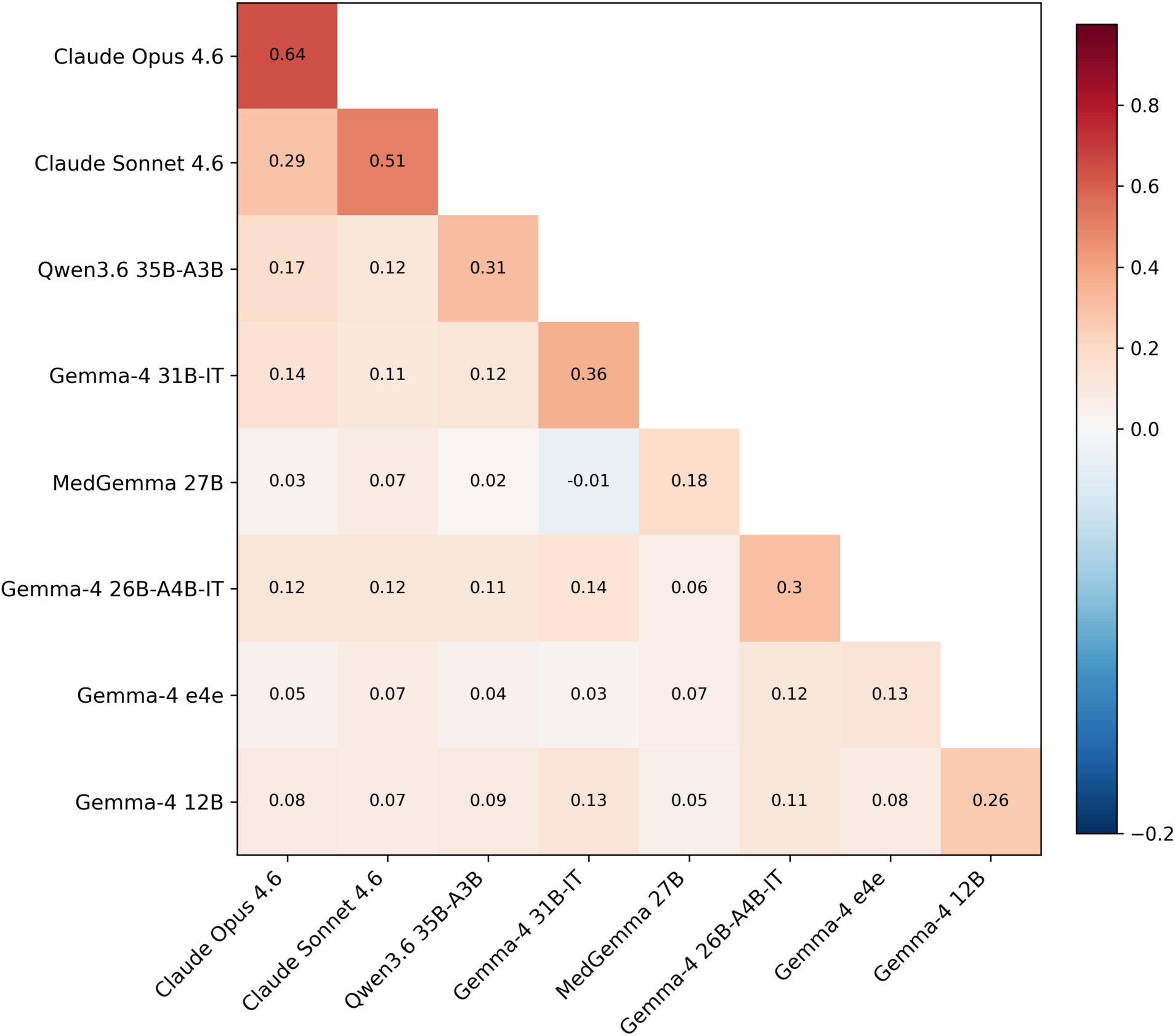
Model response correlation, computed as the task-averaged Cohen’s *κ* across the four evaluation tasks (T1-frames sliding, T1-mmode sliding, T2 lung rockets and consolidation, T3 PLAPS). Each cell is the unweighted mean of the per-task *κ* values. Within each task, off-diagonal cells average over the 5 × 5 = 25 cross-pairs of runs from the two models; diagonal cells report self-consistency, averaging over the C(5, 2) = 10 within-model run pairs. *κ* subtracts chance agreement explicitly, which keeps cells comparable across the 3-class T1 task and the binary T2/T3 tasks and across T3’s high-prevalence regime. The divergent palette accommodates the small negative cells that arise when two raters disagree more than chance would predict. Rows and columns are ordered by hierarchical clustering of the agreement values. Inter-model agreement is uniformly close to chance, well below per-model self-consistency, indicating that the models retain distinct error patterns.

The lowest self-consistency value belongs to Gemma-4-e4e (0.13); this model’s repeated runs are only marginally more consistent with each other than with predictions from any other model in the benchmark, which is consistent with elevated stochastic variability. Taken together, these observations indicate that the eight models do not produce equivalent predictions and that inter-model agreement on this benchmark cannot be explained by a shared decision boundary alone, which leaves open the possibility of complementary use through adjudication or ensembling. A direct ensemble benefit was not measured here and remains future work.

## 4 Discussion and Conclusion

The three tasks delineate current vision-language model capabilities in LUS, with this distinction driven more by input structure than by the underlying clinical complexity.

Anterior morphology (T2) shows the strongest above-chance discrimination in the benchmark. On both findings the best model exceeds the chance F1 of an all-positive predictor by approximately 0.15 and lies 9.3 and 11.2 standard deviations above the label-shuffle null (Table F1), the largest separation of any task. This standardized distance from chance is comparable across tasks, unlike raw F1, so the T2 value is a genuine cross-task maximum, whereas T1 exceeds chance by a smaller margin (4.5 standard deviations) at a low absolute F1 and T3 does not exceed chance. Both signs are visible in a single frame, a comet-tail artifact or a hepatized, tissue-like region, so uniform frame sampling preserved the discriminative evidence and the balanced cohort makes the resulting F1 (0.721 for consolidation, 0.657 for lung rockets/B-lines) a measure of discrimination rather than prevalence. This is the benchmark’s strongest evidence that VLMs can characterize clinically meaningful lung-ultrasound morphology. Between-model differences were secondary to this task-level pattern, as the benchmark primarily assesses which findings are recoverable rather than which model performs best. A single leading model was supported for lung rockets/B-lines but not for consolidation (Section 3.3, Table G1), and the model ordering should therefore be interpreted only where the paired tests agree.

The failure cases are informative in contrasting ways. Pleural sliding (T1) failed because it is a motion sign that our sparse-frame and synthetic M-mode inputs recover only partially (Section 3.2). This is a representation problem, not a sign that the models are incapable in principle. PLAPS (T3) underperformed for a different reason. As shown in Section 3.4, the prevalence-saturated posterior cohort lets models achieve high F1 by predicting positive, so the score is a dataset-balance warning rather than evidence of specificity. Furthermore, the models demonstrated only near-chance agreement with one another, well below their own self-consistency, indicating that their errors are not interchangeable. Further studies may be able to assess adjudication or ensembling, which in this study we did not directly test.

Three key conclusions emerged from this study. First, static-frame VLMs produce discriminative labels for anterior morphological findings, supporting a morphology-assist role under the input conditions tested, but not clinical deployment. Second, motion-dependent signs such as pleural sliding remain unreliable under current input representations. More expressive temporal representations than synthetic M-mode will likely be needed before sliding assessment is clinically reliable. Third, high scores on imbalanced tasks such as PLAPS must be treated as a dataset-balance warning rather than as evidence of clinical competence. Altogether, these results argue that VLM-based lung-ultrasound interpretation should be framed as a morphology-assist capability rather than a screening tool ready for clinical deployment. The most valuable next steps are expert-adjudicated ground-truth labels, balanced posterior cohorts with substantially more negative cases, repeated-run reproducibility reporting for every model, and input representations that preserve the clinically relevant motion only partially recovered by single frames and synthetic M-mode.

## 5 Limitations of the Study and Future Work

A first limitation is structural: current vision-language models do not ingest a lung-ultrasound clip as a continuous video, and neither does our pipeline. We reduce each clip to 10 uniformly sampled frames (or 10 synthetic M-mode strips), and this sparsity is not merely a design choice but a reflection of how contemporary VLMs consume moving images. Even models with native video support operate on a heavily down-sampled representation. Qwen2.5-VL processes video through dynamic-resolution encoding and absolute-time alignment over sampled frames rather than at full acquisition frame rate (Bai et al. 2025), and Gemini 1.5 samples long video at roughly one frame per second before tokenization (Gemini Team, Google 2024). The Gemma 4 family processes videos as sequences of frames, with the model card specifying support for up to sixty seconds at one frame per second (Gemma Team, Google 2026), placing it in the same low-temporal-resolution regime as the native-video models above rather than in a categorically distinct image-first regime. The practical consequence is that fine-grained, high-frame-rate motion, the precise signal that lung sliding depends on, is attenuated or discarded before the model begins to reason. Synthetic M-mode rendering, which encodes temporal change as a spatial pattern, in the manner of clinical M-mode, is a partial countermeasure in our pipeline, but it cannot fully restore the temporal resolution lost during sampling, and the residual difficulty on the sliding task is consistent with this bottleneck.

The natural remedy for weak sign recognition is domain fine-tuning, but our data make leakage-safe fine-tuning difficult. The clips are de-identified, publicly sourced atlas videos, and we cannot guarantee statistical independence between them. Frames drawn from the same clip are near-duplicates, and multiple clips may originate from the same study or patient. Under a naive split, highly correlated frames would land on both sides of the train/test boundary and produce information leakage, the “double-dipping” failure in which a model is effectively evaluated on data it has already seen. This effect is well documented in medical imaging: Tampu et al. show that splitting correlated image slices improperly inflates test accuracy by up to 0.43 in Matthews correlation coefficient (roughly 5–30% in raw accuracy) (Tampu, Eklund, and Haj-Hosseini 2022). Because de-identification strips the patient- and study-level identifiers needed to enforce strictly grouped splits, we cannot currently certify that a fine-tuning corpus would be free of such leakage. We therefore restricted this study to zero-shot evaluation. Although zero-shot evaluation avoids train/test leakage introduced during fine-tuning, pretraining-time exposure cannot be excluded.

Several dataset-level constraints further bound the conclusions. The absent-sliding class is represented by only a handful of cases, so the sliding results should be read as evidence of difficulty rather than a precise estimate of negative-class performance. The posterior PLAPS task is prevalence-saturated (see Section 3.4), so its high F1 values cannot be read as evidence of specificity. The benchmark also draws on curated educational atlas material rather than consecutive clinical acquisitions, so image quality and case mix may not reflect bedside conditions. The POCUS Atlas additionally combines an adult lung source with a smaller pediatric junior-lung subset; we report combined-cohort estimates as the primary results and provide a descriptive per-source decomposition in Appendix D, but the pediatric subset (5-21 cases per panel) is too small to support formal between-group inference.

Future work follows directly from these limitations. On the modeling side, higher-frame-rate or video-native ingestion, paired with representations that preserve temporal dynamics rather than collapsing them, would test whether the sliding bottleneck is fundamental or an artifact of sparse sampling. On the data side, assembling a grouped, leakage-controlled corpus that retains clip- and patient-level grouping while remaining de-identified would unlock safe fine-tuning and enable a fair comparison between zero-shot and adapted models. Expanding the rare negative classes and adding prospectively collected bedside clips would also strengthen the external validity of the benchmark.

## Data Availability

The benchmark dataset and associated annotations used in this study are publicly available at https://huggingface.co/datasets/bcm-liuzlab/pocus-atlas-bench. The source code, inference and evaluation pipelines, and materials supporting reproduction of the analyses are available at https://github.com/jeonglab-bcm/POCUS_RESEARCH. The underlying lung ultrasound cases were obtained from the publicly accessible POCUS Atlas at https://www.thepocusatlas.com.

## Data and Code Availability

The original lung-ultrasound cases and associated metadata are publicly available from the POCUS Atlas (https://www.thepocusatlas.com/). The derived benchmark dataset, including case annotations and task assignments, is available from Hugging Face at https://huggingface.co/datasets/bcm-liuzlab/pocus-atlas-bench. The source code used for data processing, model evaluation, and statistical analysis is available at https://github.com/jeonglab-bcm/POCUS_RESEARCH/.

## Acknowledgments

We thank the POCUS Atlas project (https://www.thepocusatlas.com) for providing open-access LUS cases with clinical annotations. Compute resources for VLM inference were provided via Amazon Web Services (AWS) Bedrock. This work was supported by the Cancer Prevention and Research Institute of Texas (CPRIT; RP240131), the Chan Zuckerberg Initiative (2023-332162), the National Institutes of Health (NIH; U54NS093793), the Eunice Kennedy Shriver National Institute of Child Health and Human Development of the NIH (P50HD103555), the NIH Division of Program Coordination, Planning, and Strategic Initiatives (DPCPSI; OT2OD040565), the Chao Endowment, the Huffington Foundation, and the Jan and Dan Duncan Neurological Research Institute at Texas Children’s Hospital.

## Footnotes

1 https://www.thepocusatlas.com/

2 The token lung slid matches both *lung slide* and *lung sliding* (and their inflections), so the two spellings need not be enumerated separately.

## References

1. Alasmawi, Hussain, Numan Saeed, and Mohammad Yaqub. 2025. “FETAL-GAUGE: A Benchmark for Assessing Vision-Language Models in Fetal Ultrasound.” arXiv Preprint. 10.48550/arXiv.2512.22278.

2. Bai, Shuai, Keqin Chen, Xuejing Liu, Jialin Wang, Wenbin Ge, Sibo Song, Kai Dang, et al. 2025. “Qwen2.5-VL Technical Report.” arXiv Preprint. 10.48550/arXiv.2502.13923.

3. Born, Jannis, Gabriel Brändle, Manuel Cossio, Marion Disdier, Nina Wiedemann, Andreas Buhre, Bastian Rieck, and Karsten Borgwardt. 2020. “POCOVID-Net: Automatic Detection of COVID-19 from a New Lung Ultrasound Imaging Dataset (POCUS).” arXiv Preprint. 10.48550/arXiv.2004.12084.

4. Gemini Team, Google. 2024. “Gemini 1.5: Unlocking Multimodal Understanding Across Millions of Tokens of Context.” arXiv Preprint. 10.48550/arXiv.2403.05530.

5. Gemma Team, Google. 2026. “Gemma 4 Model Card.” https://ai.google.dev/gemma/docs/core/model_card_4.

6. Kumar, Andre, Pawan Nandakishore, Andrea June Gordon, Evan Baum, Jai Madhok, Youyou Duanmu, and John Kugler. 2026. “Creation of an Open-Access Lung POCUS Image Database for Deep Learning and Neural Network Applications.” POCUS Journal 11 (1). 10.24908/pocusj.v11i01.19439.

7. Le, Anjie, Henan Liu, Yue Wang, Zhenyu Liu, Rongkun Zhu, Taohan Weng, Jinze Yu, et al. 2026. “U2-BENCH: Benchmarking Large Vision-Language Models on Ultrasound Understanding.” In International Conference on Learning Representations (ICLR). 10.48550/arXiv.2505.17779.

8. Li, Kunchang, Yali Wang, Yinan He, Yizhuo Li, Yi Wang, Yi Liu, Zun Wang, et al. 2024. “MVBench: A Comprehensive Multi-Modal Video Understanding Benchmark.” In Proceedings of the IEEE/CVF Conference on Computer Vision and Pattern Recognition (CVPR), 22195–206. 10.1109/CVPR52733.2024.02095.

9. Lichtenstein, Daniel A., and Gilbert A. Mezière. 2008. “Relevance of Lung Ultrasound in the Diagnosis of Acute Respiratory Failure: The BLUE Protocol.” Chest 134 (1): 117–25. 10.1378/chest.07-2800.

10. Roy, Subhankar, Willi Menapace, Sebastiaan Oei, Ben Luijten, Enrico Fini, Cristiano Saltori, Iris Huijben, et al. 2020. “Deep Learning for Classification and Localization of COVID-19 Markers in Point-of-Care Lung Ultrasound.” IEEE Transactions on Medical Imaging 39 (8): 2676–87. 10.1109/TMI.2020.2994459.

11. Sandig, Jan, Erik Küng, Lukas Aichhorn, Bernhard Schwaberger, Mathias Klemme, Natascha Wagner, Christoph Bührer, and Fabian Sternal. 2025. “Pleura-ABCDE—a Structured Expert-Based Protocol for Neonatal Lung Ultrasound Documentation and Interpretation.” The Ultrasound Journal 17: 48. 10.1186/s13089-025-00442-4.

12. Tampu, Iulian Emil, Anders Eklund, and Neda Haj-Hosseini. 2022. “Inflation of Test Accuracy Due to Data Leakage in Deep Learning-Based Classification of OCT Images.” Scientific Data 9 (1): 580. 10.1038/s41597-022-01618-6.

13. Tang, Yunlong, Jing Bi, Siting Xu, Luchuan Song, Susan Liang, Teng Wang, Daoan Zhang, et al. 2024. “Video Understanding with Large Language Models: A Survey.” IEEE Transactions on Circuits and Systems for Video Technology. 10.1109/TCSVT.2024.3483389.

14. Volpicelli, Giovanni, Mahmoud Elbarbary, Michael Blaivas, Daniel A. Lichtenstein, Gebhard Mathis, Andrew W. Kirkpatrick, Lawrence Melniker, et al. 2012. “International Evidence-Based Recommendations for Point-of-Care Lung Ultrasound.” Intensive Care Medicine 38 (4): 577–91. 10.1007/s00134-012-2513-4.

15. Wiedemann, Nina, Dianne de Korte-de Boer, Matthias Richter, Sjors van de Weijer, Charlotte Buhre, Franz A. M. Eggert, Sophie Aarnoudse, et al. 2025. “COVID-BLUeS – a Prospective Study on the Value of AI in Lung Ultrasound Analysis.” IEEE Journal of Biomedical and Health Informatics. 10.1109/JBHI.2025.3543686.

16. Yang, Zhengyuan, Linjie Li, Kevin Lin, Jianfeng Wang, Chung-Ching Lin, Zicheng Liu, and Lijuan Wang. 2023. “The Dawn of LMMs: Preliminary Explorations with GPT-4V(ision).” arXiv Preprint. 10.48550/arXiv.2309.17421.

