## Supplementary Materials for "Morphology, Not Motion: Benchmarking Vision-Language Models on Multi-Sign Lung Ultrasound Interpretation"

### A Complete Annotation Rule Set

The tables below give the complete regex patterns used to derive ground-truth labels from case metadata. Patterns are matched case-insensitively against the concatenation of case title, tags, and diagnostic categories.

The patterns use standard regular-expression notation, read as follows. Most text is matched literally and as a substring, so the token `lung slid` matches both *lung slide* and *lung sliding* (and their inflections). The form (a|b) matches either alternative. A trailing question mark makes the preceding character optional, so a-lines? matches both *a-line* and *a-lines*. The construct (?=.*x)(?=.*y) requires both $x$ and $y$ to appear anywhere in the text, in any order.

Table A1: Ground-truth label patterns. Labels are derived by case-insensitive regex matching against the concatenation of case title, tags, and diagnostic categories. For T1 sliding, unmatched cases are classified as *not assessed* and excluded from the benchmark.

| Label | Task | Type | Patterns |
|---|---|---|---|
| Sliding absent | T1 | 3-class | `absent lung slid`, `no lung slid`, `loss of lung slid`, `decreased lung slid`, `lack of lung slid`, `absence of lung slid`, `pneumothorax`, `stratosphere`, `barcode` |
| Sliding present | T1 | 3-class | `lung slid` (after removing negation phrases)[3], `seashore`; disease imputation: `pulmonary embolism`, `pe` (whole word, excluding *pediatric*), `pulmonary edema`, `cardiogenic`, `lung edema`[4] |
| Sliding both | T1 | 3-class | `lung point`, or co-occurrence of present + absent patterns |
| B-lines | T2 | boolean | `b-lines?`, `interstitial`, `alveolar syndrome`, `white lung`, `b-pattern`, `lung rockets?`, `light beam`, `spared`, `patchy`, `confluent`, `coalescing`, `coalesced`, `waterfall` |
| Consolidation | T2 | boolean | `consolidation`, `hepatization`, `hepatisation`, `tissue-like`, `shred sign`, `fractal sign`, `air bronchogram`, `c-lines?` |
| PLAPS | T3 | boolean | `plaps`, `pleural effusion`, `effusion`, `sinusoid`, `hemothorax`, `empyema`, `spine sign`, `jellyfish`, `(?=.*lung)(?=.*base)` |

Table A2: Auxiliary labels for anterior/posterior region classification (ANT = anterior, POS = posterior). Patterns are matched case-insensitively. Posterior assignment takes precedence; anterior is assigned only when posterior criteria are not met.

| Feature | Region | Patterns | Role & assignment rule |
|---|---|---|---|
| Normal posterior | POS | `lung curtain`, `curtain sign`, `(?=.*diaphragm)(?=.*a-lines?)`, `(?=.*diaphragm)(?=.*normal)` | Posterior marker; implies T3 negative when no PLAPS |
| Lung point | ANT | `lung point` | Anterior marker |
| A-lines | ANT | `a-pattern`, `a-lines?`, `normal lung slid`, `bat sign` | Anterior marker |
| Posterior rule | POS | PLAPS is true *or* normal posterior is true, **and** pathology is not pneumothorax | Posterior assignment takes precedence |

[3]The case title takes precedence over topical tags: when the title indicates absent sliding (e.g., *No Lung Sliding*), the label is *absent* despite an affirmative *lung sliding* tag (cases 0053 and 0125).

[4]Pulmonary embolism and pulmonary edema are A-/B-profile diseases that imply preserved lung sliding (no pneumothorax); sliding is therefore imputed as *present* when no direct sliding pattern matches.

| Feature | Region | Patterns | Role & assignment rule |
|---|---|---|---|
| Anterior rule | ANT | Consolidation without PLAPS, lung point, assessed sliding, B-lines without PLAPS, *or* A-lines | Anterior assigned only when posterior criteria are not met |

##### A.1 PLAPS Composition

Because the T3 label scores PLAPS as a single composite entity, the underlying finding is not part of the scored label. To characterize the composition of the PLAPS-positive cohort, we classified each PLAPS-positive case (using the T3 pipeline case filter) as carrying consolidation evidence, effusion evidence, both, or neither, applying the same annotation patterns as Table A1; the resulting composition is given in Table A3.

Table A3: Composition of the PLAPS-positive cohort by underlying finding (annotation-derived, not measured). Any consolidation involvement: 5 (14%); pure posterolateral consolidation: 1. The single *other* case (0084, Compression Atelectasis) carries no effusion or consolidation annotation and is assigned PLAPS through the positional `lung+base` rule; a trace effusion was confirmed on manual review. The cohort is effusion-dominant, and posterolateral consolidation is insufficiently represented (n=1) to constitute an independent label, consistent with treating PLAPS as a composite entity.

| PLAPS component (curated-field derived) | n | % |
|---|---|---|
| Effusion only | 31 | 84% |
| Consolidation only | 1 | 3% |
| Both effusion + consolidation | 4 | 11% |
| Other / undetermined | 1 | 3% |
| **Total PLAPS-positive** | **37** | **100%** |

##### A.2 Per-Task Dataset Statistics

Table A4 gives the exact per-task counts and class balance summarized visually in Fig. 1 (c).

Table A4: Dataset statistics per evaluation task. Prevalence indicates positive-class proportion for binary tasks; majority-class proportion for multi-class (T1).

| Task | $n$ | Classes | Prevalence |
|---|---|---|---|
| T1 Sliding | 37 | 3 | 51% present |
| T2 B-lines | 86 | 2 | 34% positive |
| T2 Consolidation | 86 | 2 | 40% positive |
| T3 PLAPS | 39 | 2 | 95% positive |

##### A.3 Excluded Cases

Table A5: Cases excluded from benchmarking (n=25). The Categories, Tags, and Body columns show the raw metadata available for regex-based annotation. Untagged cases had empty Tags fields, making reliable label derivation infeasible. Body text is excerpted; full vignettes are available in the companion dataset. Cases marked "Non-lung airway exam" (0002, 0118, 0136) are airway or tracheal examinations rather than lung-ultrasound views, and are therefore excluded as the wrong exam rather than a lung-imaging artifact. The atelectasis cases marked "BLUE non-target" (0134, 0141, 0149) are sonographically similar to consolidation but represent a distinct pathology outside the BLUE-protocol target set, and are excluded from the consolidation task to avoid label ambiguity.

| ID | Title | Categories | Tags | Body (excerpt) | Exclusion reason |
|---|---|---|---|---|---|
| 0002 | Snowstorm Sign in Endotracheal Intubation | other\|airway | Snowstorm sign\|airway Ultrasound | (...) cervical airway view during intubation attempt (...) | Non-lung airway exam |
| 0011 | Lung Re-expansion With Pigtail Catheter | other | pigtail\|catheter\|lung sliding\|expand | (...) return of lung sliding after pigtail drainage (...) | Procedure monitoring |
| 0024 | Lung Re-expansion | pneumothorax | pneumothorax\|ptx\|lung slide\|absent lung slide\|chest tube | (...) return of lung sliding after chest tube (...) | Procedure monitoring |
| 0044 | Lung Windows | Pulmonary\|normal | normal anatomy\|curved probe\|linear probe | (...) differences in view obtained with curved vs linear probe (...) | Probe comparison |
| 0054 | Pneumothorax: M-mode: Seashore vs Barcode | Pulmonary\|trauma\|pulmonarytrauma\|pneumothorax | pneumothorax\|lung slide\|barcode\|seashore\|trauma | (...) single-frame still image only (...) | Single-frame video |
| 0076 | Bar Code Sign - Pneumothorax | Pulmonary\|pulmonarytrauma\|pneumothorax | pneumothorax\|barcode | (...) single-frame still image only (...) | Single-frame video |
| 0088 | Lung Curtain | Pulmonary\|normal | lung curtain | (...) normal lung demonstrating the "lung curtain" sign (...) | Colorized duplicate |
| 0089 | Pleural Effusion | Pulmonary\|pleural effusion | pleural effusion\|exudative\|exudative pleural effusion | (...) single-frame still image only (...) | Single-frame video |
| 0097 | Thymus | other | (empty) | (...) thymus that can often be confused with lung pathology (...) | Non-pulmonary content |
| 0099 | Subcutaneous Emphysema | Trauma\|pulmonarytrauma\|other | EFAST\|subcutaneous emphysema\|e-lines | (...) patient fell 5m; E-lines artifact, lung not assessable (...) | Non-pulmonary content |

| ID | Title | Categories | Tags | Body (excerpt) | Exclusion reason |
|---|---|---|---|---|---|
| 0105 | A COVID-19 Patient with 3 days of Symptoms [1/3] | covid | (empty) | (...) confluent B-lines associated with COVID-19 pneumonia (...) | Untagged case |
| 0106 | A COVID-19 Patient with 3 days of Symptoms [2/3] | covid | (empty) | (...) patchy B-lines associated with pleural thickening (...) | Untagged case |
| 0107 | A COVID-19 Patient with 3 days of Symptoms [3/3] | covid | (empty) | (...) confluent B-lines, right posterior lung field (...) | Untagged case |
| 0108 | B-lines in COVID-19 Versus CHF | covid | (empty) | (...) side-by-side comparison of B-lines in COVID vs CHF (...) | Untagged case |
| 0109 | Hospitalized COVID-19+ Patient Day 9 [1/2] | covid | (empty) | (...) thickened irregular pleura, diffuse B-lines (...) | Untagged case |
| 0115 | Pleural Findings in COVID-19 | covid | (empty) | (...) scattered B-lines from irregular pleural line (...) | Untagged case |
| 0116 | Pleural Space - Colorized | colorized\|pulmonary | spine\| diaphragm\| pleural space\| a lines | (...) colorized overlay of pleural anatomy (...) | Colorized duplicate |
| 0117 | Normal Lung Sliding During FAST Exam | traumanormal\| lungnormal | lung sliding\| normal lung\| a-lines | (...) probe motion throughout clip, M-mode uninterpretable (...) | Probe motion artifact |
| 0118 | Esophageal Intubation | lungother\|airway | esophageal intubation\| pediatric intubation\| intubation | (...) intubation attempt into the esophagus of an 8 year old (...) | Non-lung airway exam |
| 0134 | Atelectasis in Neonate | lungother | lung collapse\| Atelectasis | (...) 32 weeks premature, neonatal atelectasis (...) | BLUE non-target |
| 0136 | Tracheal Stenosis | lungother | Airway\| tracheal diameter\| trachea\| trachea stenosis\| tracheal stenosis | (...) patient with stridor, airway visualized from neck (...) | Non-lung airway exam |
| 0140 | Lung Hepatization in Pneumonia | pneumonia | hepatization\| hepatization of lung | (...) diaphragm visible, anterior vs posterior unclear (...) | Ambiguous scan location |

| ID | Title | Categories | Tags | Body (excerpt) | Exclusion reason |
|---|---|---|---|---|---|
| 0141 | Acute Chest Syndrome | lungother | acute chest\| acute chest syndrome\| Sickle Cell | (...) sickle cell HbSS, coughing with lung findings (...) | BLUE non-target |
| 0144 | Hepatization of the Lung | pneumonia | hepatization\| hepatization of the lung | (...) 15-month-old, diaphragm visible, location ambiguous (...) | Ambiguous scan location |
| 0149 | Lung Atelectasis After Foreign Body Aspiration | lungother | lung\| atelectasis | (...) sudden respiratory distress, foreign body aspiration (...) | BLUE non-target |

### B Task Prompts

Each task used a two-pass design. Pass 1 sent the preprocessed images (Section 2.4) together with the task-specific reasoning prompt shown below and produced free-text clinical reasoning; Pass 2 then extracted the structured per-task labels from that reasoning trace via constrained decoding. The verbatim Pass 1 reasoning prompts are reproduced here.

**T1 · Pleural sliding — static frames (Pass 1 reasoning prompt)**

```
You are analyzing sequential frames from a lung ultrasound (LUS) video clip.

Analyze frame by frame first and then, assess whether pleural sliding is
present, absent, or both, using these signs:

**Signs of sliding PRESENT:**
- Shimmering or granular pleural line (bright, textured interface)
- Comet-tail artifacts or B-lines originating from the pleural line
- Seashore sign on M-mode (sandy granular pattern below the pleural line)
- Speckle pattern below the pleural line differs between consecutive frames

**Signs of sliding ABSENT:**
- Smooth, sharply defined, static pleural line
- Stratosphere / barcode sign on M-mode (horizontal lines only)
- A-lines that remain perfectly static and unchanged across all frames
- No variation in sub-pleural speckle between frames

**Signs of BOTH (lung point):**
- A visible transition point where sliding is present on one side and absent
on the other
- Alternating seashore and barcode patterns on M-mode
- Part of the pleural line shimmers while another segment is fixed
```

Examine the frames carefully for these signs. Describe what you observe, then state your conclusion.

**T1 · Pleural sliding — synthetic M-mode (Pass 1 reasoning prompt)**

```
You are analyzing 10 synthetic M-mode images extracted from a lung ultrasound video.
Each image corresponds to a different lateral position across the active ultrasound region (from left ~5% to right ~95%).

**Important:** Some edge positions often fall outside the active ultrasound region, producing nearly-black strips with minimal visible structure. Do NOT classify these as seashore or stratosphere - mark them as UNCLASSIFIABLE and exclude them from the majority count.

**A-line caveat:** A-lines (bright horizontal reverberation lines at regular intervals below the pleural line) can appear in BOTH seashore and stratosphere patterns. Their presence alone does NOT indicate stratosphere. Focus on the **background texture between the A-lines**: if it is granular/sandy → Seashore; if it is filled with continuous dense parallel horizontal lines with no granularity → Stratosphere.

**Step 1 - Per-position classification:**
For each of the 10 M-mode strips, classify the pattern as one of:
- **Seashore**: Horizontal parallel lines above the pleural line with a granular/sandy texture below (may include A-lines over the sandy background) → indicates pleural sliding is PRESENT at this position. Also look for: the pleural line itself appears irregular, wavy, or not perfectly straight - this waviness indicates motion and supports seashore even if the texture below is subtle.
- **Stratosphere/Barcode**: Dense, continuous parallel horizontal lines throughout both above and below the pleural line with NO granular texture anywhere, AND the pleural line is perfectly straight/smooth → indicates pleural sliding is ABSENT at this position.
- **Alternating (lung point)**: The same strip shows bands of granular texture (seashore) alternating with bands of parallel horizontal lines (stratosphere), cycling vertically (i.e., over time). This indicates a lung point at this position.
- **Unclassifiable**: Nearly black, minimal signal, or insufficient detail to determine pattern → exclude from decision.

Report your classification for each position (p00 through p09).

**Step 2 - Overall decision (excluding unclassifiable positions):**
- If at least one position is classified as **Alternating** → overall label is "both" (lung point)
```

```
- If there is a clear spatial transition - some positions classified as
seashore AND other positions classified as stratosphere (i.e., both patterns
coexist across different positions) → overall label is "both" (lung point)
- Otherwise, if the majority of classifiable positions show seashore pattern →
overall label is "present"
- Otherwise, if the majority of classifiable positions show
stratosphere/barcode pattern → overall label is "absent"

Examine each M-mode image carefully, describe what you observe at each
position, then state your per-position classifications and overall conclusion.
```

**T2 · Anterior pathology — B-lines and consolidation (Pass 1 reasoning prompt)**

```
You are analyzing sequential frames from a lung ultrasound (LUS) video clip of
an anterior lung zone.

Analyze frame by frame first and then, assess for B-lines and consolidation.

## B-lines Assessment

Assess the image for the presence or absence of B-lines (lung rockets):
- **B-lines present (lung_rockets = true)**: Hyperechoic vertical artifacts
arising from the pleural line, extending to the bottom of the screen without
fading, moving with lung sliding.
- **B-lines absent (lung_rockets = false)**: No vertical artifacts meeting
B-line criteria; A-lines (horizontal reverberation artifacts) may dominate.

If B-lines are present, classify the subtype:
- **septal**: Discrete, well-spaced B-lines with dark lung parenchyma visible
between them. Indicates thickened interlobular septa. Typically 3 or fewer
B-lines per intercostal space, each clearly separated.
- **ground_glass**: Confluent or coalescing B-lines that merge into a diffuse
white sheet obscuring A-lines. The lung surface appears uniformly bright.
Indicates alveolar edema or diffuse interstitial disease.
- **mixed**: Both septal (discrete, spaced) and ground_glass (confluent,
coalescing) patterns are visible in different regions or at different time
points in the clip.

## Consolidation Assessment

Assess the image for the presence of alveolar consolidation in the anterior
zone:
- **Tissue-like hepatization**: Lung parenchyma appears solid and echogenic,
resembling liver texture (hepatized), with loss of normal aeration artifacts.
- **Shred sign**: Irregular, shredded deep border between consolidated and
aerated lung.
```

```
- **Air bronchograms**: Punctate or linear hyperechoic foci within
consolidated (hepatized) lung, representing air-filled bronchi.

**Classification:**
- **consolidation = true**: One or more of the above consolidation signs are
present.
- **consolidation = false**: No consolidation signs; lung parenchyma appears
normally aerated.

If consolidation is present, classify the predominant type:
- **consolidation_type = "hepatization"**: Predominantly tissue-like
appearance with liver-like echogenicity.
- **consolidation_type = "shred_sign"**: Predominantly irregular, shredded
border pattern.
- **consolidation_type = "air_bronchogram"**: Predominantly punctate or linear
hyperechoic foci within hepatized lung.
- **consolidation_type = null**: When consolidation is false.

Describe what you observe for both B-lines and consolidation, then state your
conclusions.
```

**T3 · Posterior PLAPS (Pass 1 reasoning prompt)**

```
You are analyzing sequential frames from a lung ultrasound (LUS) video clip of
a posterior lung zone (PLAPS point - Posterolateral Alveolar and/or Pleural
Syndrome).

Assess the image for the presence of a PLAPS pattern - pleural effusion and/or
alveolar consolidation in the posterior/lateral dependent lung zone:

**Effusion signs:**
- **Quad sign**: Anechoic (black) space bounded by pleural line superiorly,
lung line inferiorly, and rib shadows laterally, forming a quadrilateral.
- **Sinusoid sign**: Cyclical movement of the lung line toward the pleural
line with respiration within a fluid collection (dynamic; best seen in
M-mode).
- **Jellyfish sign**: Floating, undulating atelectatic lung within a
surrounding effusion, resembling a jellyfish in water.

**Consolidation signs:**
- **Tissue-like hepatization**: Lung parenchyma appears solid and echogenic,
resembling liver texture (hepatized), with loss of normal aeration artifacts.
- **Shred sign**: Irregular, shredded deep border between consolidated and
aerated lung.
- **Air bronchograms**: Punctate or linear hyperechoic foci within
consolidated (hepatized) lung, representing air-filled bronchi.
```

```
**Classification:**
- **plaps = true**: One or more of the above signs are present.
- **plaps = false**: No effusion or consolidation signs; the posterior zone
appears normal with A-lines or normal lung sliding only.
- **type = "effusion"**: Predominantly fluid collection signs (quad sign,
sinusoid sign, jellyfish sign).
- **type = "consolidation"**: Predominantly tissue-like/hepatized lung without
significant free fluid.
- **type = "both"**: Both effusion and consolidation are visible.
- **type = null**: When plaps is false.

Describe what you observe, then state your conclusion.
```

### C Models

#### C.1 Model Identifiers

Table C1: Models used in the benchmark. Provider denotes the inference endpoint through which the model was invoked (not the model author): AWS Bedrock for the Claude models, OpenRouter for the three hosted Gemma/Qwen variants, and a locally-served GGUF endpoint (loaded from HuggingFace weights via an OpenAI-compatible inference server) for the three quantized Gemma/MedGemma variants. Approximate parameter counts are reported as published by the model provider; Claude parameter counts are not publicly disclosed. All models were evaluated over $N = 5$ independent complete runs per task.

| Label | Provider | Full ID | Approx. params (B) |
|---|---|---|---|
| Gemma-4 e4e | Local (HF, GGUF Q4_0) | `google/gemma-4-E4B-it-qat-q4_0-gguf:Q4_0` | 4 |
| Gemma-4 12B | Local (HF, GGUF QAT) | `unsloth/gemma-4-12B-it-qat-GGUF` | 12 |
| Gemma-4 26B-A4B-IT | OpenRouter | `google/gemma-4-26b-a4b-it` | 26 |
| MedGemma 27B | Local (HF, GGUF Q8_0) | `unsloth/medgemma-27b-it-GGUF:Q8_0` | 27 |
| Gemma-4 31B-IT | OpenRouter | `google/gemma-4-31b-it` | 31 |
| Qwen3.6 35B-A3B | OpenRouter | `qwen/qwen3.6-35b-a3b` | 35 |
| Claude Sonnet 4.6 | AWS Bedrock | `us.anthropic.claude-sonnet-4-6` | undisclosed |
| Claude Opus 4.6 | AWS Bedrock | `us.anthropic.claude-opus-4-6-v1` | undisclosed |

### D Per-Source Decomposition: Adult and Junior Subsets

#### D.1 Rationale and Procedure

The main benchmark (Section 3) aggregates over the full evaluable cohort, which can mask differences between the adult lung atlas and the pediatric junior atlas. Table D1 reports the same per-task F1 computed separately on each subset. These per-source metrics are obtained by matching the ground truth to the case-level source annotation and re-aggregating the existing model outputs (no re-inference) under the case-level bootstrap of Section 2. Fig. D1 visualizes the four panels in which the junior subset is informative. Because the junior subset is small in every task, we treat this decomposition as descriptive and perform no formal between-group testing.

Table D1: Per-(panel, model) F1 on the adult and junior subsets, with case-level 95% bootstrap confidence intervals computed under the resampling procedure described in Section 2. Junior subsets contain $n = 6$ cases for T1 panels, $n = 21$ for T2 panels, and $n = 5$ for T3 PLAPS, so the junior intervals are wide and, for several cells, span most of the [0, 1] range; the table is descriptive disclosure of partition-specific behavior, not a basis for between-group inference. [a] The T3 PLAPS junior subset (5 cases, high positive-class prevalence) yields F1 $\approx$ 1.00 for most models; the row is retained for completeness but T3 is omitted from Fig. D1, as a 5-case binary subset is uninformative for visual comparison across models.

| Panel | Model | Adult F1 [95% CI] (n) | Junior F1 [95% CI] (n) |
|---|---|---|---|
| T1 Frames — Pleura Sliding Macro F1 | claude_opus-4.6 | 0.27 [0.14–0.38] (31) | 0.28 [0.00–0.55] (6) |
| | claude_sonnet-4.6 | 0.44 [0.24–0.62] (31) | 0.28 [0.00–0.56] (6) |
| | gemma-4-12b | 0.17 [0.07–0.27] (31) | 0.17 [0.00–0.27] (6) |
| | gemma-4-26b-a4b-it | 0.40 [0.19–0.61] (31) | 0.41 [0.10–0.63] (6) |
| | gemma-4-31b-it | 0.30 [0.17–0.45] (31) | 0.10 [0.00–0.22] (6) |
| | gemma-4-e4e | 0.34 [0.18–0.50] (31) | 0.11 [0.00–0.25] (6) |
| | medgemma-27b | 0.27 [0.12–0.41] (31) | 0.19 [0.00–0.30] (6) |
| | qwen3.6-35b-a3b | 0.21 [0.11–0.32] (31) | 0.40 [0.09–0.62] (6) |
| T1 M-mode — Pleura Sliding Macro F1 | claude_opus-4.6 | 0.46 [0.28–0.65] (31) | 0.52 [0.10–0.87] (6) |
| | claude_sonnet-4.6 | 0.40 [0.28–0.51] (31) | 0.40 [0.10–0.63] (6) |
| | gemma-4-12b | 0.21 [0.11–0.32] (31) | 0.17 [0.00–0.27] (6) |
| | gemma-4-26b-a4b-it | 0.46 [0.20–0.67] (31) | 0.39 [0.11–0.63] (6) |
| | gemma-4-31b-it | 0.36 [0.23–0.47] (31) | 0.36 [0.00–0.58] (6) |
| | gemma-4-e4e | 0.11 [0.02–0.20] (31) | 0.27 [0.00–0.52] (6) |
| | medgemma-27b | 0.34 [0.19–0.51] (31) | 0.13 [0.00–0.29] (6) |
| | qwen3.6-35b-a3b | 0.23 [0.11–0.35] (31) | 0.22 [0.00–0.33] (6) |
| T2-1 Lung Rockets — F1 | claude_opus-4.6 | 0.47 [0.31–0.61] (65) | 0.42 [0.12–0.67] (21) |
| | claude_sonnet-4.6 | 0.45 [0.30–0.57] (65) | 0.40 [0.11–0.64] (21) |
| | gemma-4-12b | 0.56 [0.41–0.69] (65) | 0.48 [0.19–0.71] (21) |
| | gemma-4-26b-a4b-it | 0.61 [0.45–0.73] (65) | 0.56 [0.24–0.80] (21) |
| | gemma-4-31b-it | 0.67 [0.51–0.79] (65) | 0.71 [0.36–0.94] (21) |
| | gemma-4-e4e | 0.47 [0.32–0.60] (65) | 0.42 [0.17–0.64] (21) |
| | medgemma-27b | 0.30 [0.14–0.44] (65) | 0.46 [0.00–0.75] (21) |
| | qwen3.6-35b-a3b | 0.57 [0.40–0.71] (65) | 0.50 [0.15–0.78] (21) |
| T2-2 Consolidation — F1 | claude_opus-4.6 | 0.55 [0.35–0.70] (65) | 0.64 [0.35–0.83] (21) |
| | claude_sonnet-4.6 | 0.51 [0.29–0.69] (65) | 0.70 [0.42–0.90] (21) |
| | gemma-4-12b | 0.52 [0.31–0.68] (65) | 0.64 [0.35–0.85] (21) |
| | gemma-4-26b-a4b-it | 0.53 [0.31–0.71] (65) | 0.59 [0.29–0.82] (21) |
| | gemma-4-31b-it | 0.57 [0.37–0.72] (65) | 0.74 [0.46–0.92] (21) |
| | gemma-4-e4e | 0.30 [0.11–0.48] (65) | 0.37 [0.00–0.64] (21) |
| | medgemma-27b | 0.15 [0.00–0.35] (65) | 0.17 [0.00–0.44] (21) |
| | qwen3.6-35b-a3b | 0.76 [0.58–0.88] (65) | 0.56 [0.22–0.80] (21) |
| T3 PLAPS — F1 [a] | claude_opus-4.6 | 0.97 [0.92–1.00] (34) | 1.00 [1.00–1.00] (5) |
| | claude_sonnet-4.6 | 0.97 [0.92–1.00] (34) | 1.00 [1.00–1.00] (5) |
| | gemma-4-12b | 0.97 [0.92–1.00] (34) | 1.00 [1.00–1.00] (5) |
| | gemma-4-26b-a4b-it | 0.97 [0.92–1.00] (34) | 1.00 [1.00–1.00] (5) |
| | gemma-4-31b-it | 0.95 [0.89–1.00] (34) | 1.00 [1.00–1.00] (5) |
| | gemma-4-e4e | 0.83 [0.72–0.92] (34) | 0.89 [0.57–1.00] (5) |
| | medgemma-27b | 0.69 [0.52–0.82] (34) | 0.75 [0.33–1.00] (5) |
| | qwen3.6-35b-a3b | 0.97 [0.92–1.00] (34) | 1.00 [1.00–1.00] (5) |

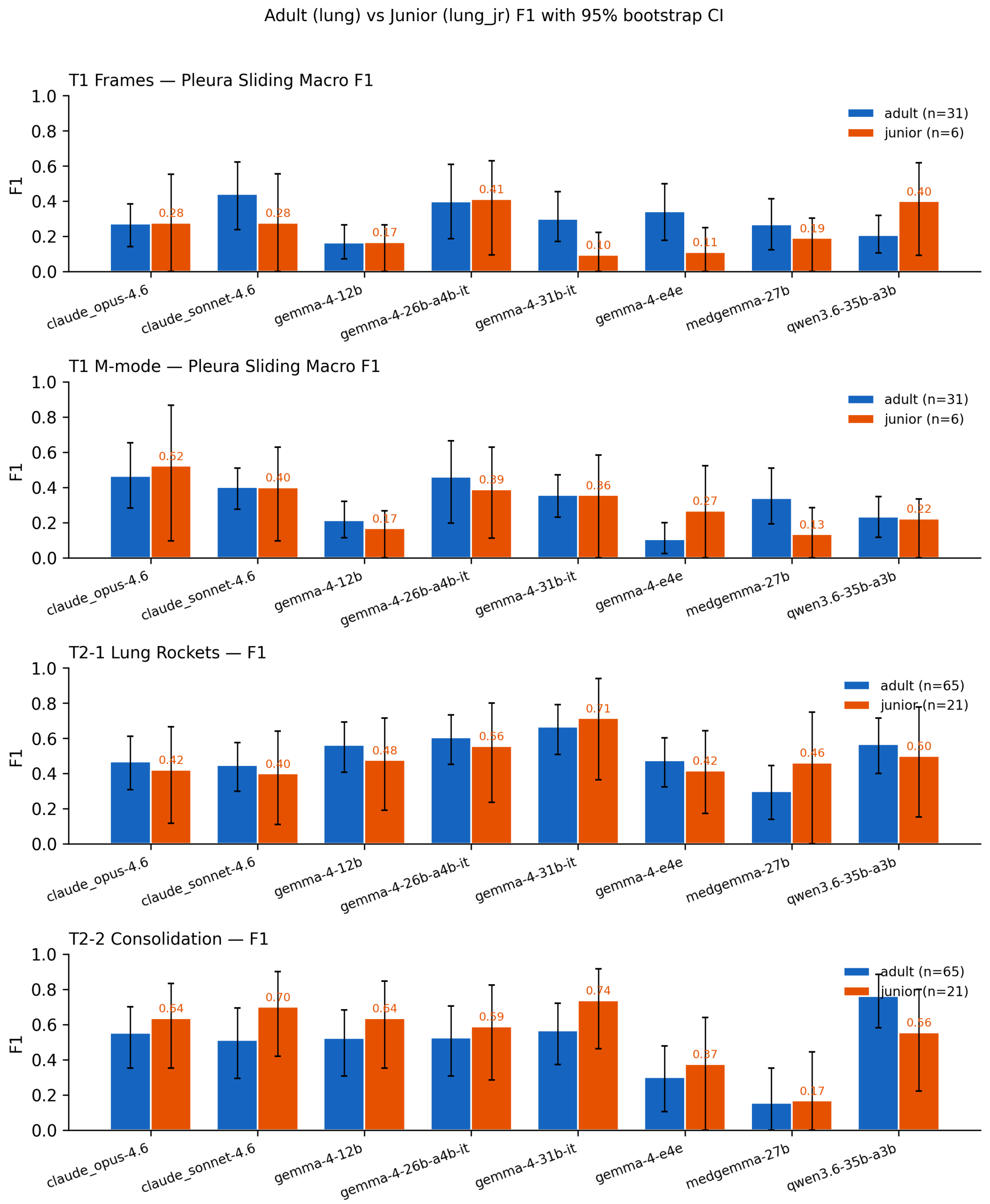


Figure D1: Per-source F1 with 95% bootstrap confidence intervals on the T1 Frames, T1 M-mode, T2-1 Lung Rockets, and T2-2 Consolidation panels. Adult subsets contain 31 cases for T1 panels and 65 for T2 panels; junior subsets contain 6 and 21 respectively. T3 PLAPS is omitted for the reason stated in Table D1. Combined-cohort estimates and the headline benchmark figure remain in Section 3.

##### D.2 Interpretation

The combined-cohort estimates in Section 3 remain the primary quantitative claim of this paper; the per-source view is supplied for transparency and to bound the inferences a reader can draw from the aggregate numbers. With junior sample sizes ranging from 5-21 across panels, the case-level bootstrap intervals on the junior column are wide enough that for most (panel, model) cells the adult and junior intervals overlap substantially. We therefore describe the decomposition as observational and refrain from characterizing any model as differentially robust to the pediatric subset on the basis of this sample alone.

Two patterns appear at the level of point estimates and may merit follow-up once a larger pediatric corpus is available. First, on the T1 frame-based sliding panel, 2 open-weight Gemma variants exhibit a substantial point-estimate drop from adult to junior (`gemma-4-31b-it`: 0.30 to 0.10; `gemma-4-e4e`: 0.34 to 0.11), while `qwen3.6-35b-a3b` moves in the opposite direction (0.21 to 0.40); none of these shifts are individually significant given the junior interval widths, but the directional inconsistency across architectures is itself informative. Second, on T2-2 consolidation, several models score higher on the junior than on the adult subset (e.g., `claude_sonnet-4.6`: 0.51 to 0.70; `gemma-4-31b-it`: 0.57 to 0.74), a pattern more consistent with partition-specific positive-class prevalence or labeling distribution than with a uniform pediatric generalization deficit. We have not adjusted these observations for multiple comparisons across the eight models and five panels, and they should not be read as significant on this sample.

### E BLUE Protocol Mapping

Fig. E1 shows the BLUE protocol's sequential assessment and highlights the sign-recognition nodes evaluated by our benchmark, color-coded by task: blue nodes denote T1 (sliding detection), green nodes T2 (anterior-zone pathology), and orange nodes T3 (posterolateral PLAPS). T1, T2, and T3 collectively cover all lung sign-level questions in the protocol; only the integration step (combining signs into a profile and pathology diagnosis) and the venous DVT assessment (a lower-extremity vascular scan separate from lung imaging) remain outside scope.

### F Above-Chance Discrimination

The per-task above-chance verdicts are stated in the Results sections (Section 3.2, Section 3.3, Section 3.4). This appendix consolidates the five tests into a single reference table so the observed F1, the chance baseline, and the standardized effect size can be compared side by side.

To confirm that reported F1 reflects genuine discrimination rather than a prevalence artifact, we compared the best-performing model on each task against a label-shuffle null (Section 2). The ground-truth labels were permuted within each complete run, the mean F1 across runs was recomputed, and this was repeated 10,000 times to build the null distribution. Table F1 reports the observed mean F1, the chance F1 of a no-information predictor (an all-positive predictor, $2p/(1+p)$, for the binary tasks and the majority-class predictor macro-F1 for the multi-class T1 task), and a standardized effect size $z$, the number of null standard deviations separating the observed F1 from the mean of the shuffle distribution.

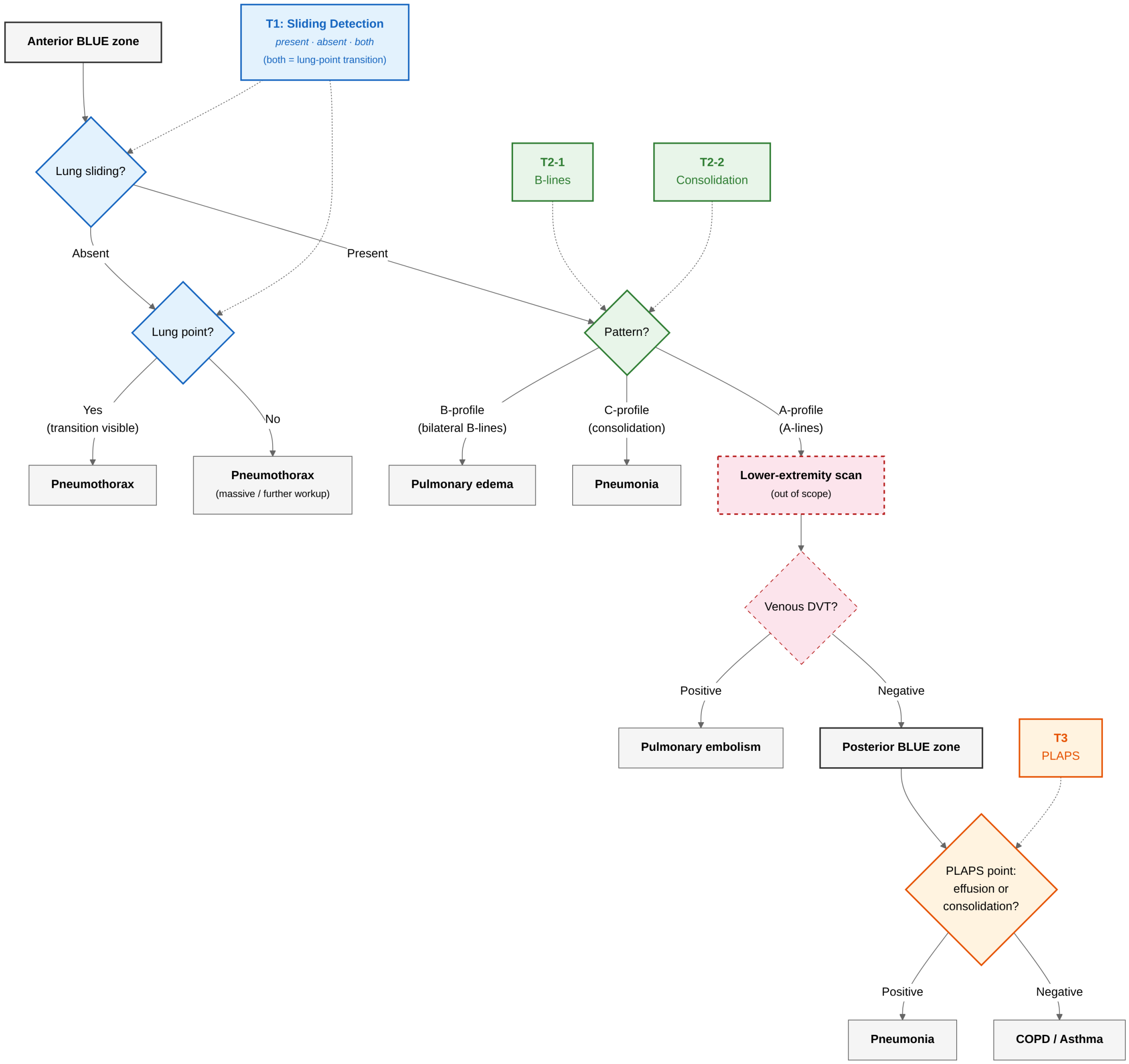


Figure E1: BLUE protocol decision tree (adapted from Lichtenstein & Mezière 2008) with benchmark task mapping. The anterior and posterior labels denote the BLUE protocol's assessment zones, not a recorded probe position; per-case zone assignment is inferred from annotation markers (see Section 2.2). Node colors mark the sign-recognition tasks (T1/T2/T3); dashed-red elements denote the lower-extremity DVT scan, which lies outside benchmark scope.

Table F1: Above-chance discrimination per task. $z$ is the standardized distance of the observed F1 from the label-shuffle null (number of null standard deviations); a larger $z$ means the result is further above chance. For T3 the null has zero variance, because an all-positive predictor scores the same F1 under any label permutation on a 95%-positive cohort, so $z$ is undefined and the observed 0.974 equals the chance F1 exactly.

| Task | Best model | Metric | Observed F1 | Chance F1 | $z$ |
|---|---|---|---|---|---|
| T1 Frames | Gemma 4 26B A4B | macro-F1 | 0.425 | 0.226 | 4.5 |
| T1 M-mode | Claude Opus 4.6 | macro-F1 | 0.486 | 0.226 | 4.5 |
| T2 B-lines | Gemma 4 31B | binary-F1 | 0.657 | 0.504 | 9.3 |
| T2 Consolidation | Gemma 4 31B | binary-F1 | 0.721 | 0.567 | 11.2 |
| T3 PLAPS | Claude Opus 4.6 | binary-F1 | 0.974 | 0.974 | n/a |

T2 sits furthest above chance ($z$ of 9.3 and 11.2 on the two findings), well beyond T1 ($z$ of 4.5 on both variants) even though T1's raw margin above its chance baseline is similar, because the balanced T2 cohort gives a tighter null. T1 is clearly above chance but at a low absolute F1. T3 is not above chance at all. Shuffling the labels does not change the F1 of an all-positive predictor on a 95%-positive cohort, so the observed 0.974 is indistinguishable from the no-information baseline.

### G Between-Model Comparison on T2

The T2 between-model verdicts are stated in the T2 Results section (Section 3.3). This appendix reports the underlying paired-test statistics for the two highest-mean-F1 models on each anterior finding. As defined in Section 2, a difference is treated as established only when both the pooled exact McNemar test and the paired t-test on the five per-run F1 values reject at the 0.05 level.

Table G1: Paired between-model comparison on T2. Model A is the top mean-F1 model and Model B the next. $b$ and $c$ are the pooled discordant-pair counts (cases where one model is correct and the other is wrong). Paired $t$ is the paired t-test statistic on the five per-run F1 values and Paired $t$ $p$ its two-sided p-value. A difference is treated as established only when both the pooled exact McNemar test and the paired t-test on the five per-run F1 values reject at the 0.05 level. The lung-rocket lead is established (both tests reject), whereas the consolidation lead is not (neither test rejects), so the consolidation ranking is reported descriptively.

| Task | Model A | Model B | Mean F1 A | Mean F1 B | McNemar $b$ / $c$ | McNemar $p$ | Paired $t$ | Paired $t$ $p$ |
|---|---|---|---|---|---|---|---|---|
| Lung rockets/B-lines | Gemma 4 31B | Gemma 4 12B | 0.657 | 0.584 | 104 / 41 | $1.7 \times 10^{-7}$ | 4.32 | 0.012 |
| Consol-idation | Gemma 4 31B | Qwen3.6 35B A3B | 0.721 | 0.623 | 63 / 48 | 0.184 | 1.97 | 0.120 |

The per-run McNemar p-values are 0.014, 0.061, 0.052, 0.248, and 0.001 for lung rockets/B-lines, and 0.230, 0.308, 0.481, 0.332, and 0.248 for consolidation. Only the lung-rocket comparison combines a significant pooled test with a significant paired t-test, which is why the manuscript asserts a single leading model for lung rockets/B-lines but not for consolidation.
